# Expanding the genetics and phenotypes of ocular congenital cranial dysinnervation disorders

**DOI:** 10.1101/2024.03.22.24304594

**Authors:** Julie A. Jurgens, Brenda J. Barry, Wai-Man Chan, Sarah MacKinnon, Mary C. Whitman, Paola M. Matos Ruiz, Brandon M. Pratt, Eleina M. England, Lynn Pais, Gabrielle Lemire, Emily Groopman, Carmen Glaze, Kathryn A. Russell, Moriel Singer-Berk, Silvio Alessandro Di Gioia, Arthur S. Lee, Caroline Andrews, Sherin Shaaban, Megan M. Wirth, Sarah Bekele, Melissa Toffoloni, Victoria R. Bradford, Emma E. Foster, Lindsay Berube, Cristina Rivera-Quiles, Fiona M. Mensching, Alba Sanchis-Juan, Jack M. Fu, Isaac Wong, Xuefang Zhao, Michael W. Wilson, Ben Weisburd, Monkol Lek, Ocular CCDD Phenotyping Consortium, Harrison Brand, Michael E. Talkowski, Daniel G. MacArthur, Anne O’Donnell-Luria, Caroline D. Robson, David G. Hunter, Elizabeth C. Engle

**Author notes:** Correspondence: Elizabeth Engle, MD, Boston Children’s Hospital.

## Abstract

**Purpose:** To identify genetic etiologies and genotype/phenotype associations for unsolved ocular congenital cranial dysinnervation disorders (oCCDDs).

**Methods:** We coupled phenotyping with exome or genome sequencing of 467 pedigrees with genetically unsolved oCCDDs, integrating analyses of pedigrees, human and animal model phenotypes, and *de novo* variants to identify rare candidate single nucleotide variants, insertion/deletions, and structural variants disrupting protein-coding regions. Prioritized variants were classified for pathogenicity and evaluated for genotype/phenotype correlations.

**Results:** Analyses elucidated phenotypic subgroups, identified pathogenic/likely pathogenic variant(s) in 43/467 probands (9.2%), and prioritized variants of uncertain significance in 70/467 additional probands (15.0%). These included known and novel variants in established oCCDD genes, genes associated with syndromes that sometimes include oCCDDs (e.g., *MYH10, KIF21B, TGFBR2, TUBB6),* genes that fit the syndromic component of the phenotype but had no prior oCCDD association (e.g., *CDK13, TGFB2*), genes with no reported association with oCCDDs or the syndromic phenotypes (e.g., *TUBA4A, KIF5C, CTNNA1, KLB, FGF21*), and genes associated with oCCDD phenocopies that had resulted in misdiagnoses.

**Conclusion:** This study suggests that unsolved oCCDDs are clinically and genetically heterogeneous disorders often overlapping other Mendelian conditions and nominates many candidates for future replication and functional studies.

## INTRODUCTION

Ocular congenital cranial dysinnervation disorders (oCCDDs) are rare neurogenic disorders that present as limited extraocular movement in one or multiple directions of gaze and/or ptosis. Extraocular muscles move the eyes and eyelids and are innervated by three cranial nerves (CN) that originate from brainstem motor nuclei: the oculomotor (CN3), trochlear (CN4), and abducens (CN6). oCCDDs result from defects in the development of these ocular motor neurons and/or their axons (Supplementary Fig. 1) and can occur in isolation or together with syndromic phenotypes.^1^ CN3-oCCDDs include congenital fibrosis of the extraocular muscles (CFEOM), congenital ptosis (ptosis), and Marcus Gunn jaw-winking syndrome (MGJWS). CFEOM is defined by non-progressive upgaze limitation, typically with ptosis and variable limitation of downgaze and horizontal gaze. Ptosis can also occur in isolation, or together with eyelid opening in response to jaw movement in MGJWS (MGJWS(+)ptosis).^2^ Rarely, MGJWS occurs without ptosis (MGJWS(-)ptosis), resulting in eye-opening with jaw movement. Inverse MGJWS (INV-MGJWS(-)ptosis) refers to eyelid closure upon jaw movement. CN4-oCCDDs include fourth nerve palsy (CN4-palsy), characterized by the inability to adduct and depress the eye, and Brown syndrome, which presents as limited elevation in adduction and can alternatively result from non-CN4-related mechanical restriction. CN6-oCCDDs include Duane retraction syndrome (DRS), congenital sixth-nerve palsy (CN6-palsy), and horizontal gaze palsy. DRS is characterized by limited ocular abduction and narrowing of the palpebral fissure with globe retraction on attempted adduction. CN6-palsy presents as limited abduction, and horizontal gaze palsy as limited abduction and adduction, both in the absence of globe retraction. Atypical oCCDDs that do not fit cleanly into these categories are referred to as “CCDD-not otherwise specified’’ (CCDD-NOS).

Multiple oCCDD genes have been reported (Fig. 1A), as have genes underlying conditions that, rarely, are misdiagnosed as oCCDDs, including myasthenias, myopathies, or extraocular muscle maldevelopment syndromes.^3,4,5,6^ Most oCCDD genes were identified through analysis of large pedigrees and/or homogenous endophenotypes with shared etiologies. This leaves a large, genetically unsolved cohort of predominantly small, phenotypically heterogeneous pedigrees. To identify genetic causes and genotype/phenotype associations in this cohort, we performed exome or genome sequencing and prioritized single nucleotide variants (SNVs), small insertions/deletions (indels), and structural variants (SVs) to detect known and novel candidate protein-coding oCCDD etiologies.

**Figure 1.**
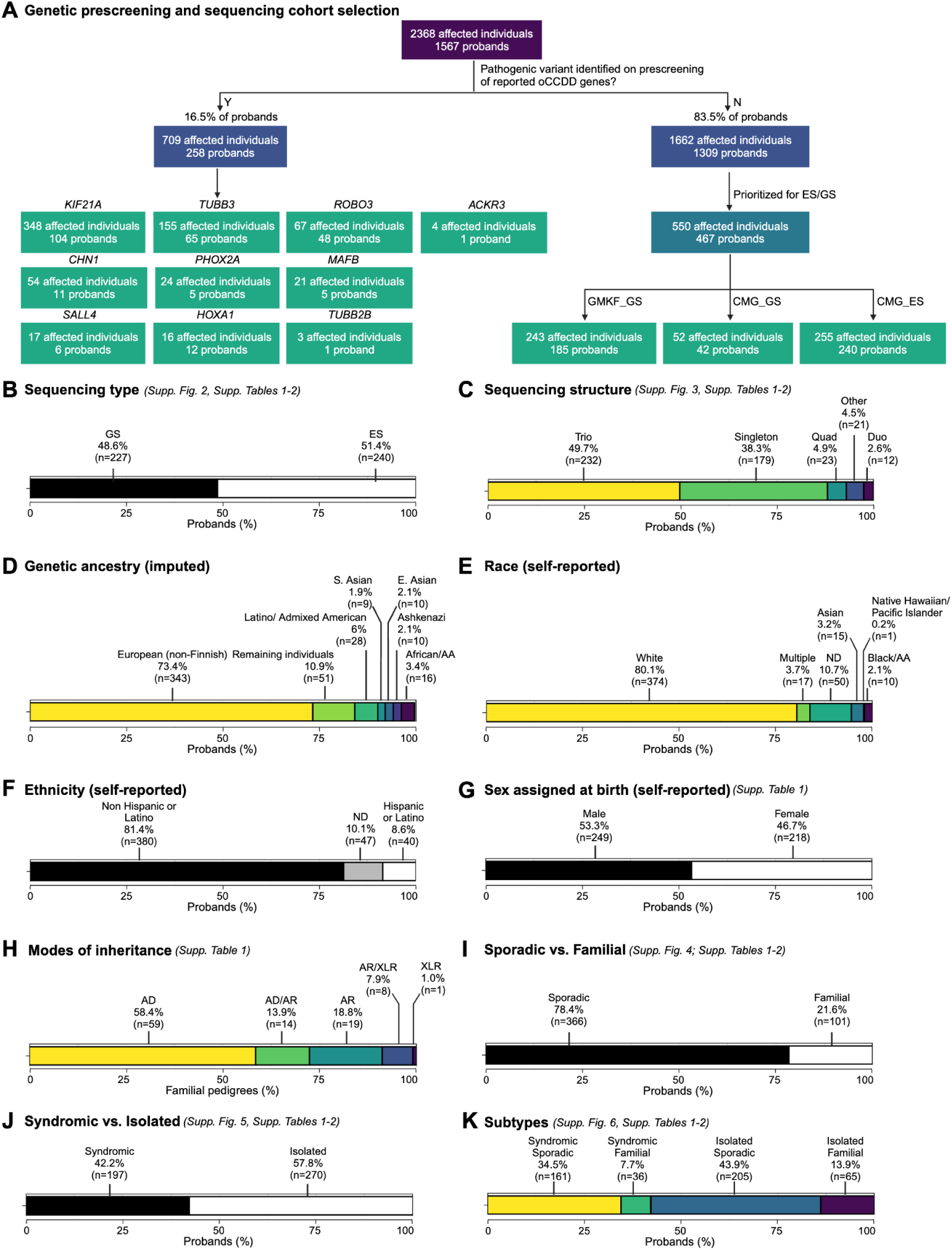
Selection and composition of sequenced probands in the ocular CCDD cohort. **A:** A large initial cohort of individuals with ocular CCDDs (oCCDDs) and their relatives were enrolled in our research study at Boston Children’s Hospital from August 1992 to June 2019. Individuals who had pathogenic variants in reported oCCDD genes identified on pre-screening were not included in the study (left side). The remaining individuals (right side) were prioritized for exome/genome sequencing if they screened negative for known common genetic etiologies of oCCDDs, had sufficient DNA quality and quantity, consented to broad genomic data sharing, and had additional syndromic features. Sequencing was performed through GMKF (GS) and the Broad CMG (GS and ES). **B-K:** Categorization of the proportions of 467 probands in the sequenced oCCDD cohort according to various metrics, as follows: B: Sequenced by GS versus ES. C: Sequenced as singletons, duos, trios, quads, or other (>4 total members of the pedigree sequenced). D: Imputed genetic ancestry groups. E: Self-reported race. F: Self-reported ethnicity. G: Self-reported sex assigned at birth. H: Modes of inheritance. I: Sporadic vs. familial. J: Syndromic vs. isolated. K: Both syndromic and sporadic, syndromic and familial, isolated and sporadic, or isolated and familial. For all relevant panels, accompanying supplementary figures and tables are denoted. Abbreviations: AA=African American, AD=autosomal dominant, AR=autosomal recessive, CCDD=congenital cranial dysinnervation disorder, CMG=Centers for Mendelian Genomics, E.=East, ES=exome sequencing, GMKF=Gabriella Miller Kids First, GS=genome sequencing, ND=not described, oCCDD=ocular congenital cranial dysinnervation disorder, S.=South, XLR=X-linked recessive.

## MATERIALS AND METHODS

The following sections are expanded upon in Supplementary Methods.

### Cohort enrollment, data collection, and phenotyping

We studied 467 genetically unresolved probands encompassing 11 oCCDDs (CFEOM, ptosis, MGJWS(+)ptosis, MGJWS(-)ptosis, INV-MGJWS(-)ptosis, CN4-palsy, Brown syndrome, DRS, CN6-palsy, horizontal gaze palsy, or CCDD-NOS) and their relatives (Supplementary Methods). Most probands (403/467, 86.3%) were pre-screened for pathogenic variants in reported oCCDD genes (Fig. 1A).

Demographics were collected via survey and self-reported by participants or their parents/legal guardians. Phenotypic data were obtained through retrospective review of clinical records, questionnaires, and updates from participants and their clinicians. Ocular motility data were reviewed by pediatric ophthalmologists, orthoptists, and neurologists (authors DGH, MCW, SM, ECE). Affected individuals were assessed for non-oculomotor syndromic phenotypes in 20 categories (Supplementary Methods). Brain magnetic resonance images (MRIs) were reviewed by a pediatric neuroradiologist (author CR) for image quality and for abnormalities of cranial nerves, extraocular muscles, structural brain, and other non-brain structures.

Individuals with an oCCDD and at least one major or two minor congenital anomalies were categorized as syndromic, while participants not meeting those criteria were categorized as isolated. Probands with or without a known family history of oCCDDs were designated as familial or sporadic. Co-occurring defects analysis determined whether oCCDDs and syndromic phenotypes co-occurred more frequently than by chance.^7^

### DNA sequencing

Exome/genome sequencing and genetic ancestry imputation were performed on 467 genetically unsolved pedigrees (550 affected and 1108 total individuals).

### SNV/indel analysis and prioritization

SNVs/indels were filtered using inheritance models, allele frequency, quality control, and variant annotation. Trios and larger pedigrees were assessed for frameshifting and non-frameshifting indels and for missense, nonsense, or splice site-altering SNVs in protein-coding genes. Singletons and duos were assessed only for variants in known oCCDD genes, strong candidate genes, genes mutated in >1 proband, and variants annotated as pathogenic/ likely pathogenic in ClinVar^8^ in additional genes. SNV/indels were prioritized using analyses of known biology and genotype/phenotype associations, animal models,^9^ statistical and pathway analyses of *de novo* variants (DNVs),^10,11^ and AlphaMissense scores^12^.

### SV analysis and prioritization

SVs perturbing coding sequences were identified, jointly genotyped, and annotated using GATK-SV (https://github.com/broadinstitute/gatk-sv) or GATK-gCNV and filtered for allele frequency, inheritance models, quality control, and variant annotation.^13,14^

### Interpretation of variant pathogenicity and submission to ClinVar

Using recommendations from the American College of Genetics and Genomics and Association for Molecular Pathology (ACMG/AMP)^15^ and the Clinical Genome Resource (ClinGen), SNVs/indels and SVs were prioritized and classified as pathogenic (P), likely pathogenic (LP), or uncertain significance (VUS), and submitted to ClinVar (Data Availability).^16^ Classified variants were grouped into five categories (Supplementary Methods).

### Genotype/phenotype correlations

For pedigrees with highlighted genetic findings, clinical features were reported using Human Phenotype Ontology (HPO)^17^ and assessed for genotype/phenotype associations.

## RESULTS

### Definition of the oCCDD cohort

At the onset of this project, we had enrolled 1567 pedigrees with oCCDDs; the phenotype was familial in 364 (23.4%). A genetic etiology had been identified for 258 pedigrees (16.5%), which were not included in this study (Fig. 1A, Supplementary Results).

Among 1309 unsolved pedigrees, 467 were sequenced (550 affected and 1108 total individuals). Pedigrees were eligible for exome/genome sequencing if they screened negative for common genetic etiologies, had sufficient DNA quality and quantity, and consented to data sharing in controlled access repositories. Pedigrees with familial oCCDDs and/or syndromic features were further prioritized. 227 (48.6%) and 240 (51.4%) pedigrees had genome and exome sequencing, respectively. Most were sequenced as trios (49.7%) or singletons (38.3%), with fewer duos (2.6%), quads (4.9%), or “other” pedigrees (>4 individuals; 4.5%). Five pedigrees for which we reported a genetic etiology during the study were included (Pedigrees 38, 48, 56, 98, 144).^18–22^ Clinical genetic testing before exome/genome analysis explained the syndromic phenotypes of two unrelated individuals (proband ENG_AKL and affected individual 178_04). Genetic imputation revealed European ancestry in 73.4% of probands; this was corroborated by self-reported race and ethnicity (reported at a cohort-wide level). Male-assigned sex at birth and consanguinity were reported in 53.0% and 4.3% of probands, respectively. The oCCDD was familial in 101 pedigrees (21.6%), of which 58.4% displayed autosomal dominant inheritance. 270/467 probands (57.8%) had an isolated oCCDD, and almost half had both isolated and sporadic oCCDDs (43.9%). See Fig. 1A-K, Supplementary Results, Supplementary Figs. 2-6, Supplementary Tables 1-2.

DRS was the most common oCCDD in the cohort (n=198, 42.4%). oCCDDs were unilateral (60.6%), bilateral (27.0%), or of unknown laterality (12.4%), and synkinesis was described in 60.8%. Syndromic findings were noted in 197 probands (42.2%). We identified co-occurring phenotypes underlying known syndromes but no distinguishable novel syndromes. See Fig. 2A-D, Supplementary Results, Supplementary Figures 7-8, Supplementary Tables 1-4.

**Figure 2.**
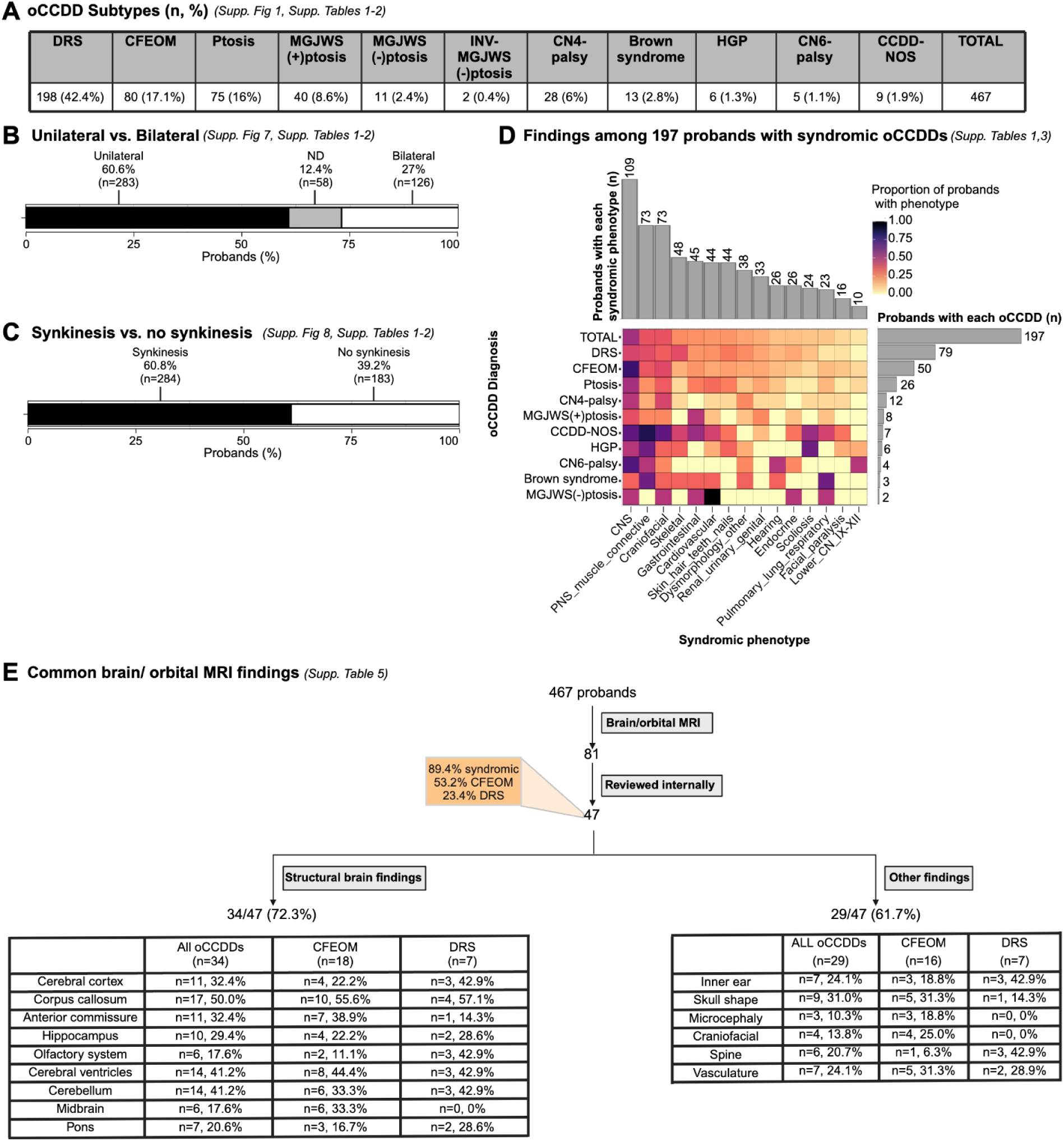
Phenotypes among sequenced probands in the ocular CCDD cohort. **A:** Numbers and percentages of probands with each oCCDD subtype. **B:** oCCDD laterality among all oCCDD probands. **C:** Evaluation of synkinesis among all oCCDD probands. D: Syndromic findings among the 197 probands with syndromic oCCDDs. Proportions of probands with each syndromic finding are represented colorimetrically within the heatmap. Gray bars on the right side of the heatmap show the total number of syndromic probands with each oCCDD subtype (i.e. 79 probands had DRS), while gray bars above the heatmap show the total number of probands with involvement in the corresponding syndromic category beneath the heatmap (e.g., 109 total probands had CNS involvement). **E:** Brain/orbital MRI findings among oCCDD probands. Of the 467 sequenced oCCDD probands, 81 had clinically obtained MRIs, 47 of which were available for review (of whom 89.4% had a syndromic oCCDD). Proportions of individuals with various structural brain anomalies and other findings are provided. For all relevant panels, accompanying supplementary figures and tables are denoted. Abbreviations: CCDD=congenital cranial dysinnervation disorder, CCDD-NOS=CCDD not otherwise specified, CFEOM=congenital fibrosis of the extraocular muscles, CMG=Centers for Mendelian Genomics, CN4-palsy=fourth nerve palsy, CN6-palsy=sixth nerve palsy, CNS=central nervous system, DRS=Duane retraction syndrome, E.=East, ES=exome sequencing, GMKF=Gabriella Miller Kids First, GS=genome sequencing, HGP=horizontal gaze palsy, INV-MGJWS(-)ptosis=inverse Marcus Gunn jaw-winking synkinesis without congenital ptosis, MGJWS(+)ptosis=Marcus Gunn jaw-winking synkinesis with congenital ptosis, MGJWS(-)ptosis=Marcus Gunn jaw-winking synkinesis without congenital ptosis, ND=not described, PNS=peripheral nervous system, Ptosis=congenital ptosis, S.=South, XLR=X-linked recessive.

### Brain and Orbital MRI findings

Brain/orbital MRIs for 47 probands were available for review. Of these probands, 53.2% had CFEOM, 23.4% had DRS, and 89.4% were syndromic. Thirteen scans were optimized for cranial nerve/extraocular muscle detection, and all had ocular cranial nerve and/or extraocular muscle abnormalities. In most, the affected cranial nerves/extraocular muscles on MRI were consistent with the clinically diagnosed oCCDD, though occasional images revealed phenocopies such as extraocular muscle-tethering orbital bands (Fig. 2E, Supplementary Results, Supplementary Table 5).

Scans revealed structural brain and non-brain anomalies in 34/47 (72.3%) and 29/47 (61.7%) of probands, respectively. While some have been reported with oCCDDs (e.g., inner ear findings),^3^ others may have been previously underrecognized (e.g., vascular anomalies; Fig. 2E, Supplementary Results, Supplementary Table 5).

### SNV/indel analyses

Among all 467 pedigrees, SNV/indel filtering yielded 48,194 rare coding variants in 16,503 genes (Fig. 3A). Among 276 pedigrees with ≥3 individuals sequenced, the yield was 6,102 variants in 4,288 genes (Supplementary Table 6). To further prioritize these, we implemented analyses of genotype/phenotypes, animal models, and statistical and pathway analyses of DNVs.

**Figure 3.**
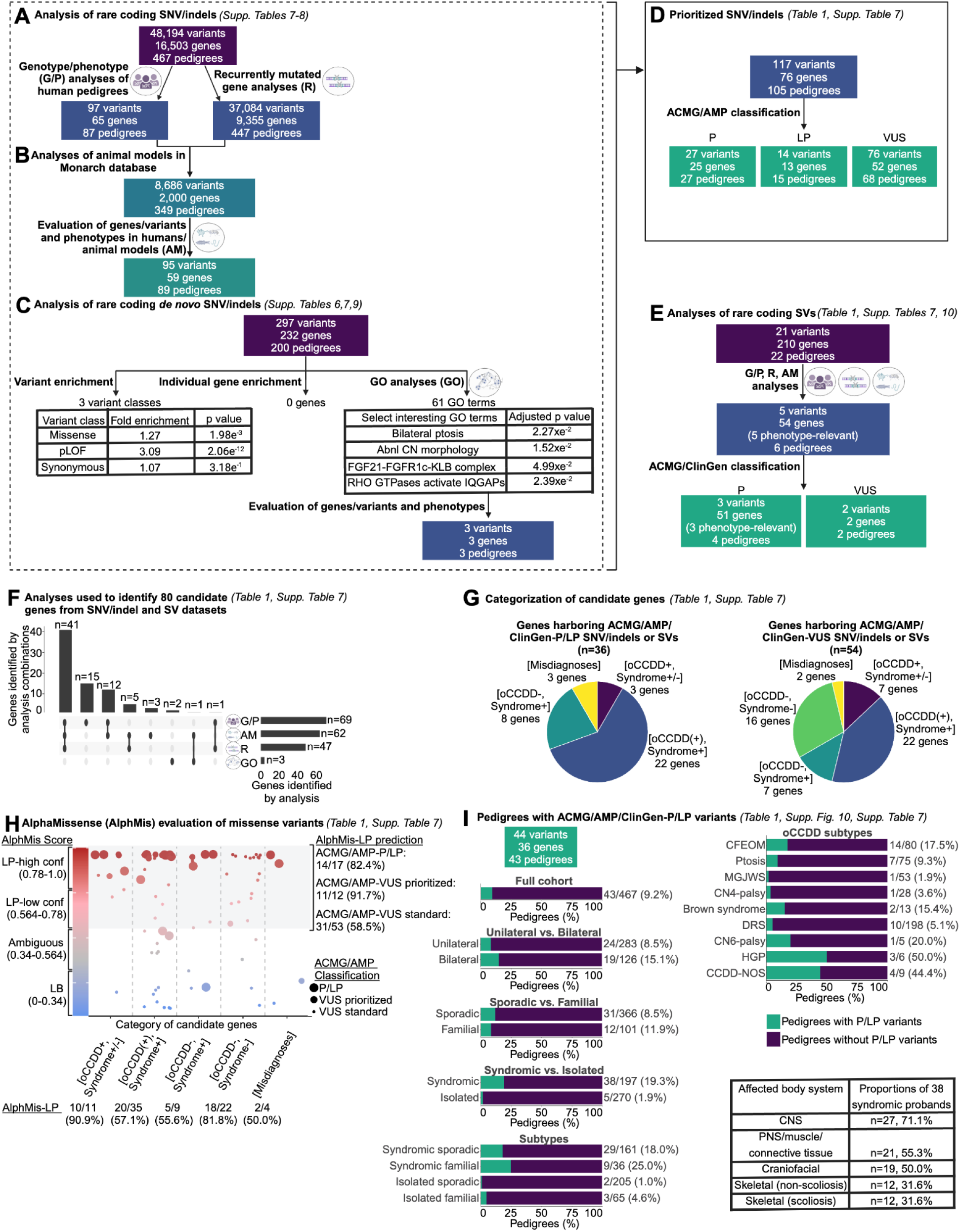
Workflow of genetic analyses. **A-C:** Workflow for analyses of rare coding SNVs and indels. A: Pedigree-based genotype/phenotypes (G/P) analyses and recurrently mutated gene analyses. Rare coding SNVs/indels in all 467 pedigrees were identified and subjected to genotype/phenotype analyses as described (Supplementary Methods, “SNV/indel filtering and prioritization” and “Biological prioritization of SNVs/indels” sections). Genes that had SNVs/indels meeting the parameters defined in Supplementary Methods, “SNV/indel filtering and prioritization” and that were mutated in more than one pedigree were defined as recurrently mutated genes. Recurrently mutated genes were queried in all 467 pedigrees regardless of specific oCCDD diagnosis. **B:** Workflow for animal model analyses (“AM” analyses). Each of the 65 genes that harbored rare coding SNVs/indels derived from G/P analyses and the 9,355 genes identified from recurrently mutated gene (“R”) analyses (from ‘A’) were annotated for putatively relevant animal model phenotypes in the Monarch database, which yielded 2,000 genes with putatively oCCDD-relevant animal models. 59 candidate genes / 95 variants were prioritized from this analysis (defined in Supplementary Methods, “Animal model analyses” section). **C:** Workflow for statistical and gene ontology (GO) analyses of *de novo* variants (DNVs). Rare coding DNV SNV/indels were assessed for overall enrichment of various classes of DNVs (left) and enrichment of DNVs in individual genes (middle) through DenovolyzeR, and for enrichment of genes in specific pathways through GO analysis (right). Numbers of total and specific classes of enriched GO terms are displayed, along with select enriched GO terms of biological interest. Genes derived from the GO terms “FGF21-FGFR1c-KLB complex” and “RHO GTPases activate IQGAPs” were nominated for ACMG/AMP classification. **D:** ACMG/AMP classification. In total from the G/P, R, AM, and/or GO analyses of SNVs/indels in (A-C), 117 variants in 76 genes were prioritized and classified by the ACMG/AMP criteria. The gene/variant counts were derived from (A-C) minus redundant genes/variants. In total, only 76 distinct genes are represented among the three ACMG/AMP classification categories, as some genes had variants in more than one classification category. **E:** Rare coding SVs were prioritized using G/P, R, and AM analyses as described for SNV/indels, which led to the nomination of 5 SVs for classification by ACMG/ClinGen criteria. **F:** UpSet plot summarizing the combinations of analyses (G/P, AM, R, and GO) used to derive the 80 candidate genes whose variants were nominated for ACMG/AMP/ClinGen classification of SNV/indels (76 genes from D) or SVs (4 genes not overlapping with SNV/indels). Vertical bars denote numbers of candidate genes identified by each combination of analyses. Horizontal bars denote numbers of genes identified by each analysis type in total (numbers were obtained by adding genes with prioritized SNV/indels plus genes with prioritized SVs minus genes represented redundantly between the SNV/indel and SV classes; Supplementary Table 7). **G:** Classification of candidate genes harboring ACMG/AMP/ClinGen-P/LP SNV/indels and SVs (left chart), or ACMG/AMP/ClinGen-VUS SNV/indels and SVs (right chart). Among the 80 genes with P/LP or VUS variants (F), 10 were represented in both categories. Genes were stratified into five categories. Purple [oCCDD+,Syndrome+/-]: genes that were definitively associated with oCCDDs before this study and were genetically pre-screened in most probands. Blue [oCCDD(+),Syndrome+]: genes that had at least occasional prior oCCDD association but were typically part of specific monogenic syndromes and thus not pre-screened. Dark green [oCCDD-,Syndrome+]: genes that fit the syndromic component of each proband’s phenotype but that, to our knowledge, have no prior oCCDD association. Light green [oCCDD-,Syndrome-]: genes that, to our knowledge, had no reported association with either the oCCDD or non-CCDD phenotype of the probands who harbor them. Yellow [Misdiagnoses]: genes associated with alternative non-neurogenic/ non-CCDD etiologies and represent misdiagnoses or oCCDD phenocopies. **H:** AlphaMissense (AlphMis) could be used to assess 82 of the ACMG/AMP classified missense SNVs. X-axis: Numbers of missense variants and percent scored as AlphMis-LP in each of the five categories as defined in **(G).** Y-axis (left side): AlphMis scores on a scale of zero to one accompanied by the corresponding pathogenicity score (LB, Ambiguous, LP-low confidence, LP-high confidence). Scores are color-coded from blue (LB) to red (LP). Each dot on the plot represents a separate missense variant, and dot sizes correspond with ACMG/AMP classifications (large dots: ACMG/AMP-P/LP variants, medium dots: strongly prioritized ACMG/AMP-VUS, small dots: standardly prioritized ACMG/AMP-VUS). Strongly prioritized ACMG/AMP-VUS are the missense VUS denoted in Table 1; while these are formally classified as ACMG/AMP-VUS, we concluded that these variants have compelling biological and/or genotype/phenotype evidence and are most likely to be substantiated over time. Standardly prioritized ACMG/AMP-VUS are all additional missense ACMG/AMP-VUS denoted in Supplementary Table 7 that we prioritized but that currently have less supportive evidence than the strongly prioritized VUS. High-confidence and low-confidence AlphMis-LP variants are encompassed by the gray shaded region of the graph and compared to their independently obtained ACMG/AMP classifications (right side of the graph); numerical summaries are provided for each, for instance: 14/17 (82.4%) ACMG/AMP-P/LP variants were also scored as LP by AlphaMissense. **I:** Rates of ACMG/AMP/ClinGen-P/LP variants (SNVs, indels, and SVs) obtained for the full cohort (left) and individual subgroups (top right). Rates are given as the number of pedigrees within each group who had ACMG/AMP/ClinGen-P/LP variant(s) relative to the total number of pedigrees within that group. Green= pedigrees with ACMG/AMP/ClinGen-P/LP variant(s); Purple= pedigrees without ACMG/AMP/ClinGen-P/LP variant(s). Among 38 syndromic probands who had ACMG/AMP/ClinGen-P/LP variant(s), the most frequently affected body systems are shown (bottom right). For all relevant panels, accompanying supplementary figures and tables are denoted. Abbreviations: AM=animal model analyses, abnl=abnormal, ACMG=American College of Genetics and Genomics, AlphMis=AlphaMissense, AMP=Association for Molecular Pathology, CCDD=congenital cranial dysinnervation disorder, CCDD-NOS=CCDD not otherwise specified, CFEOM=congenital fibrosis of the extraocular muscles, ClinGen=Clinical Genome Resource, CN=cranial nerve, CN4-palsy=fourth nerve palsy, CN6-palsy=congenital sixth nerve palsy, CNS=central nervous system, conf=confidence, DRS=Duane retraction syndrome, GO=gene ontology analyses, G/P=genotype/phenotype analyses, HGP=horizontal gaze palsy, HPO=human phenotype ontology, indel=small insertion/deletion, LB=likely benign, LP=likely pathogenic, MGJWS=Marcus Gunn jaw-winking synkinesis, misc=miscellaneous, oCCDD=ocular congenital cranial dysinnervation disorder, P=pathogenic, pLOF=predicted loss of function (nonsense, splicing, or frameshift), PNS=peripheral nervous system, ptosis=congenital ptosis, R=recurrently mutated gene analyses, SNV=single nucleotide variant, SV=structural variant, VUS=variant of uncertain significance.

We first analyzed individual pedigrees and performed genotype/phenotype correlations, prioritizing 97 variants in 65 genes among 87 pedigrees. We identified strong candidate variants among 13 pedigrees in 6 oCCDD genes (*KIF21A*, *TUBB3*, *PHOX2A, CHN1*, *MAFB*, *ROBO3*). Next, we prioritized additional genes/variants based on predictive scores, functional annotations, and reported associations with the oCCDD and/or syndromic phenotype in the corresponding pedigree (Table 1, Fig. 3A, Supplementary Results, Supplementary Table 7).

**Table 1.**
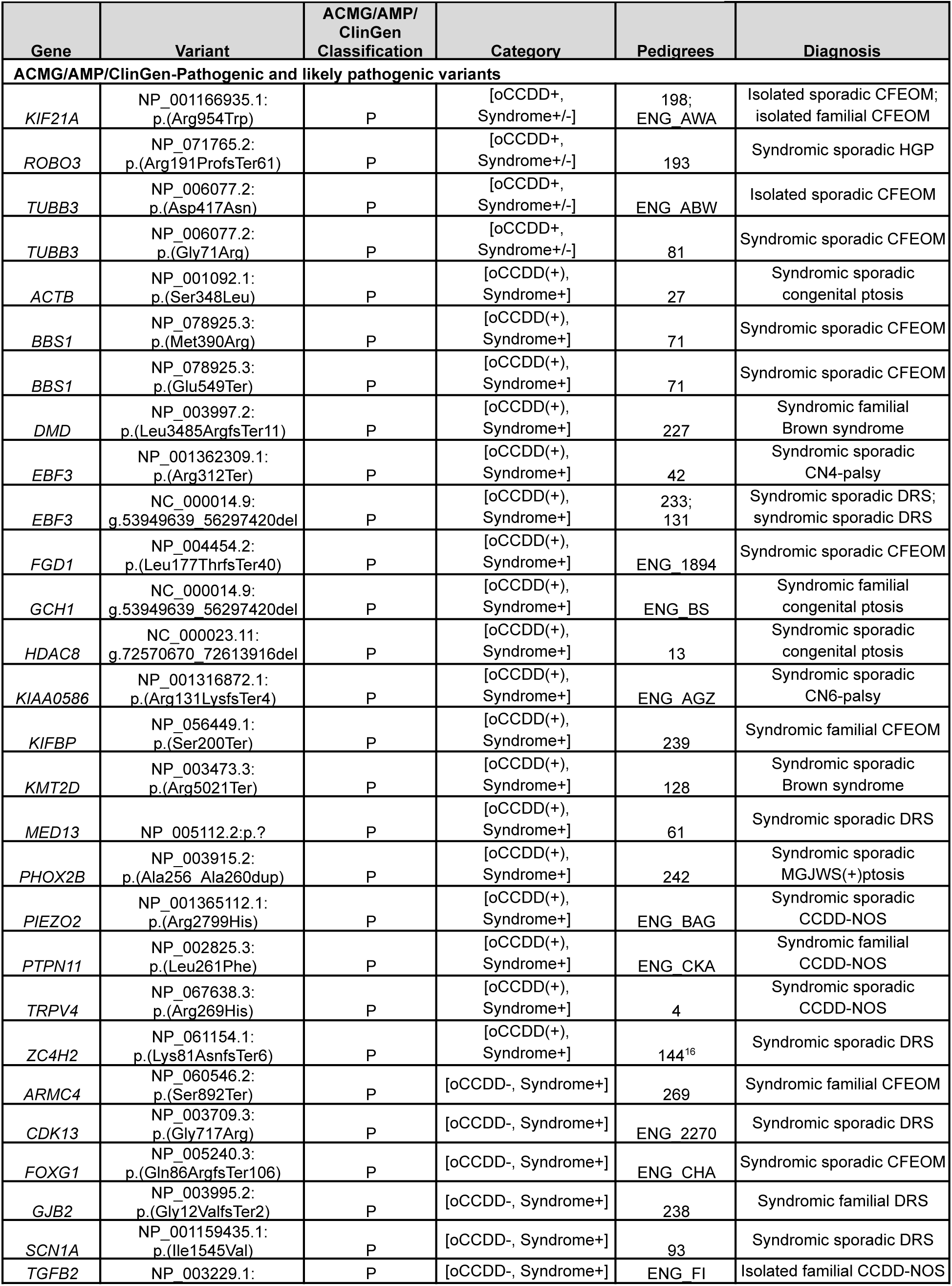
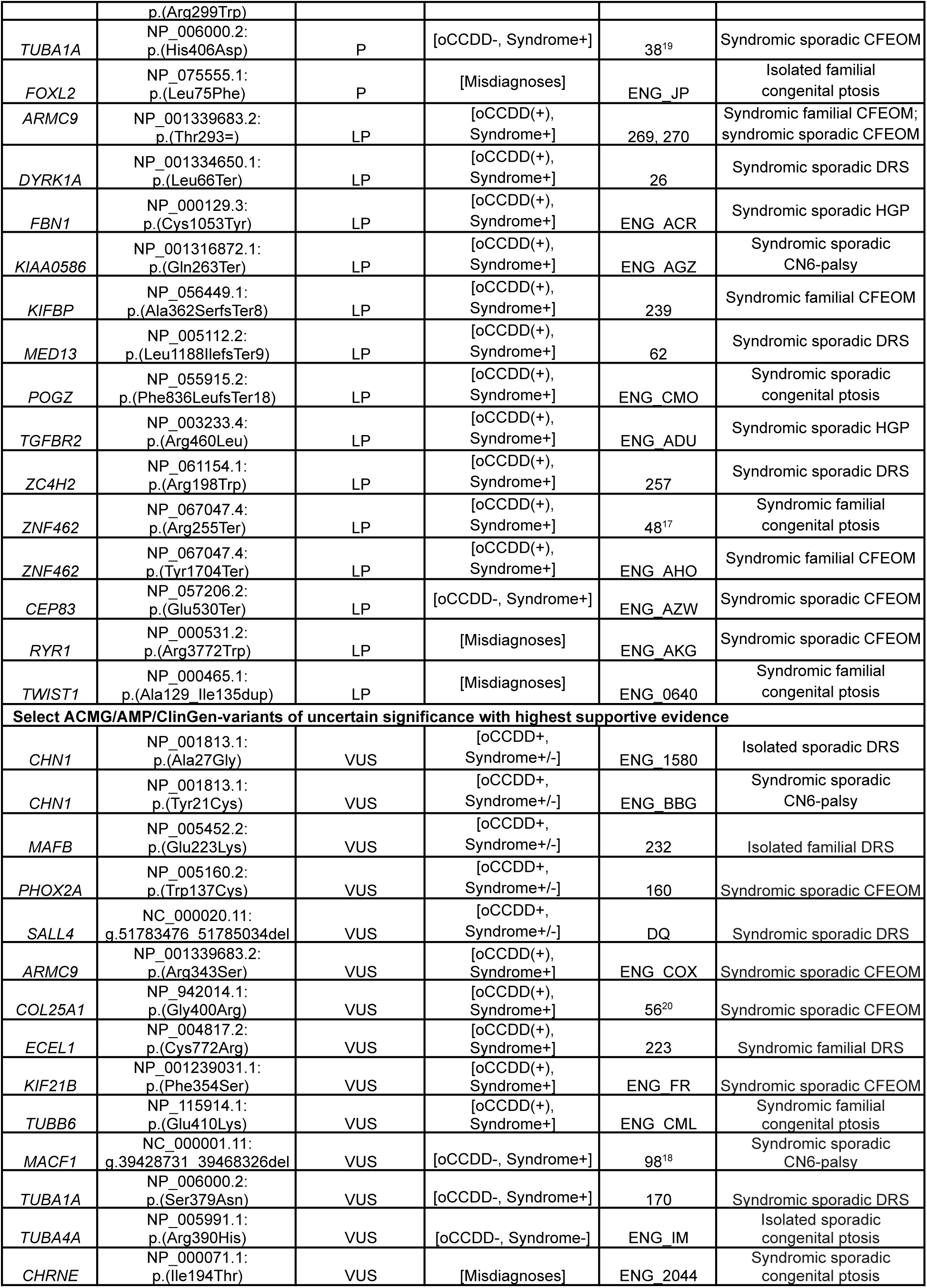
Prioritized genetic findings in the oCCDD cohort. Included are all variants identified in the oCCDD cohort that were classified as pathogenic or likely pathogenic, followed by a subset of variants of uncertain significance selected based on having the most supportive evidence of potential pathogenicity. Supplementary Table 7 provides detailed variant and phenotype data for each of these probands and for the probands who harbor additional variants described in the manuscript. Genes are delineated into five categories, defined in the main text and in Figure 3. Abbreviations: ACMG-American College of Medical Genetics and Genomics, AMP=Association for Molecular Pathology, CCDD=congenital cranial dysinnervation disorder, CCDD-NOS=CCDD not otherwise specified, CFEOM=congenital fibrosis of the extraocular muscles, ClinGen=Clinical Genome Resource, CN4-palsy=fourth nerve palsy, CN6-palsy=sixth nerve palsy, DRS=Duane retraction syndrome, HGP=horizontal gaze palsy, LP-likely pathogenic, MGJWS(+)ptosis=Marcus Gunn jaw-winking syndrome with congenital ptosis, oCCDD-ocular congenital cranial dysinnervation disorder, P-pathogenic, VUS-variant of uncertain significance.

We identified strong candidate variants in *TUBA1A, TUBB6, TUBB4A,* and *TUBB*, complementing our previous reports of tubulin-encoding *TUBB3* and *TUBB2B* as oCCDD genes.^1^ We reported *TUBA1A* as a novel CFEOM gene during this study (pedigree 38)^21^ and subsequently identified an additional variant in syndromic sporadic DRS pedigree 170. We identified a *TUBB6* p.(Glu410Lys) variant in syndromic familial ptosis pedigree ENG_CML; this substitution has been reported as P/LP for tubulinopathies in four paralogs (*TUBB2A, TUBB2B, TUBB3,* and *TUBB4A;* ClinVar Variation IDs: 986830, 1195195, 6967, 135658). Moreover, a separate *TUBB6* missense variant was reported in a 5-generation syndromic ptosis pedigree (MIM617732).^23^ Similarly, we identified a *TUBA4A* heterozygous p.(Arg390His) variant in isolated sporadic ptosis pedigree ENG_IM; this residue aligns with tubulinopathy-associated residues in four paralogs (*TUBA1A, TUBB2A, TUBB2B,* and *TUBB3;* ClinVar Variation IDs: 488628, 423490, 418531, 450183, 1214258, 160177, 1203166, 429413, 1320230, 219257). These findings support the putative pathogenicity of these missense changes. Separate missense or loss-of-function variants in *TUBA4A* have been reported in amyotrophic lateral sclerosis (MIM616208) but not with oCCDDs. Finally, we identified strong candidate variants in additional tubulin-encoding genes *TUBB4A* in isolated familial Brown syndrome and *TUBB* in isolated sporadic CFEOM (pedigrees 216, ENG_0678; Supplementary Table 7).

We identified heterozygous *MYH10* variants in five pedigrees with CN3-oCCDDs: ENG_CKM (isolated familial CFEOM), ENG_ASW (syndromic sporadic CFEOM), ENG_YY (isolated sporadic MGJWS(+)ptosis), ENG_PJ (isolated sporadic MGJWS(+)ptosis), and ENG_CGO (isolated sporadic MGJWS(-)ptosis) (Supplementary Table 7, Supplementary Figure 9). *MYH10* is highly missense-constrained within humans (missense z=5.006)^24^ and encodes a nonmuscle myosin that modulates actin dynamics. Separate *MYH10* heterozygous missense or loss-of-function variants were recently associated with neurodevelopmental phenotypes, which included ptosis in 3 individuals, lateral rectus muscle weakness in 1 individual, and CN5/CN7 palsy in 1 individual.^25,26^ Additional *MYH10* missense variants have been reported in probands with ptosis, coloboma, and craniofacial dysmorphisms (Scheidecker et al., personal communication).

Among 8 probands, we identified 12 variants in 7 genes associated with ciliopathies including Joubert syndrome (*KIAA0586, ARMC4, BBS1, CEP83, ARMC9, TOGARAM1, WDR5;* Supplementary Table 7). In Joubert syndrome, eye movement abnormalities and ptosis are common, and oCCDD-like phenotypes are reported infrequently.^27^ Our findings substantiate the association between genetically diverse ciliopathies and oCCDDs.

Finally, 4 probands harbored a variant in one of three TGF-beta pathway genes (*FBN1, TGFB2, TGFBR2*; Supplementary Table 7). These genes are associated with connective tissue disorders that are occasionally accompanied by ocular abnormalities including putative CFEOM,^28^ but these are not strongly associated oCCDD genes.

Among 447 pedigrees, we identified 9,355 recurrently mutated genes. These were prioritized if also nominated by additional analyses of genotype/phenotypes above, or of animal models or DNVs below.

For many oCCDD genes, animal models have recapitulated human oCCDD phenotypes.^1,3,4^ Thus, we queried whether strong candidate genes identified from genotype/phenotype analysis or recurrently mutated candidate genes were annotated for oCCDD-relevant animal model phenotypes in the Monarch database^9^, and further prioritized 95 variants in 59 genes among 89 pedigrees (Fig. 3B, Supplementary Results, Supplementary Table 8). We identified a homozygous *ECEL1* variant that disrupts a residue involved in disulfide bonding in pedigree 223 with familial DRS and arthrogryposis. Biallelic *ECEL1* variants are reported to cause distal arthrogryposis (MIM615065)^29^ that, rarely, is accompanied by an oCCDD. Supporting *ECEL1* variants in DRS pathogenicity, *Ecel1* mouse models have abnormal CN6 innervation (Table 1; Supplementary Tables 7-8).^30^ Animal model analyses also prioritized candidate variants in genes without known human oCCDD involvement (*KIF5C*, *NES, CUX1, GNAS, FER, ACTR1B, OLIG2,* and *SEMA3F;* Supplementary Results; Supplementary Tables 7-8). Among these, the neuronal kinesin-encoding gene *KIF5C* was mutated in three singletons: ENG_1561 (syndromic sporadic DRS), ENG_ABE (syndromic familial DRS), and ENG_UV (isolated sporadic

CCDD-NOS with intermittent blinking during smiling). *KIF5C* is highly missense-constrained within humans (missense z=4.054).^24^ Distinct *KIF5C* variants have been identified in human cortical brain malformations with variable syndromic involvement;^31^ unfortunately, our probands did not have MRIs to evaluate cortical brain malformations. Intriguingly, *Kif5c^-/-^* mice have fewer CN6 motor neurons,^32^ consistent with DRS. Moreover, downregulation of the oCCDD gene *TUBB3* increases *KIF5C* motility and cargo transport.^33^ While this is an intriguing candidate, all pedigrees with *KIF5C* variants in our cohort are singletons, and the syndromic phenotypes in the two DRS probands are disparate from one another and from known *KIF5C*-associated syndromic findings.

We next identified and performed statistical and pathway analyses of 297 DNVs in 232 genes among 200 pedigrees (Fig. 3C). Probands had an enrichment of missense and predicted loss-of-function DNVs but not synonymous DNVs (fold enrichment=1.27, 3.09, 1.07; p=1.98e^-3^, 2.06e^-12^, 3.18e^-1^; Poisson test). Because DNVs were not significantly enriched in any individual gene (Supplementary Results), we tested enrichment among genes associated with shared HPO terms, signaling pathways, or protein complexes via GO analysis. Many HPO terms were enriched (Supplementary Table 9), including terms supporting the involvement of DNV-genes in oCCDDs, such as bilateral ptosis and abnormal cranial nerve morphology (p_adj_=2.27e^-2^ and 1.52e^-2^). Additionally, we identified pathways or protein complexes that highlighted novel candidate genes. For instance, the enriched pathway term “RHO GTPases activate IQGAPs” (p_adj_=2.39e^-2^) included oCCDD genes (*TUBB3, TUBA1A, ACTB*), but also highlighted *CTNNA1*, which had a DNV in pedigree 99 with syndromic sporadic CFEOM (Supplementary Tables 7,9). *CTNNA1* variants have been associated with retinopathies (MIM116805) but not with oCCDDs. The term for the FGF21-FGFR1c-KLB protein complex was also enriched (p_adj_=4.99e^-2^), with DNVs in *FGF21* in syndromic sporadic CFEOM pedigree 91 and in *KLB* in syndromic sporadic CCDD-NOS pedigree ENG_CKP (Supplementary Tables 7,9). Notably, variants in other FGF signaling genes are reported to cause phenotypes present in pedigree 91, including hypogonadism, syndactyly, craniofacial dysmorphisms, and developmental delay.^34^ Moreover, hypogonadism has been reported in *Fgf21^-/-^*mice.^35^ Part of the syndromic phenotype in ENG_CKP is craniosynostosis, a phenotype associated with other FGF signaling genes but not yet *KLB*. Finally, a single *Klb^-/-^* mouse image demonstrates periorbital abnormalities reminiscent of ptosis or globe retraction.^36^

### ACMG/AMP classification of prioritized SNVs/indels

Through qualitative manual review of genes obtained from analysis of genotype/phenotypes, animal models, and pathways above, we selected a subset of 117 SNV/indels among 105 probands for ACMG/AMP classification (Fig. 3D), including the four probands for whom we reported a causal SNV/indel.^18,19,21,22^ For this subset, we also annotated HPO terms and performed additional extensive genotype/phenotype correlations for oCCDD and syndromic phenotypes (Supplementary Tables 1,7).

In total, ACMG/AMP classification identified 41 P/LP SNVs/indels among 39/467 probands (8.4%; Fig. 3D, Table 1, Supplementary Table 7). Three probands had compound heterozygous variants in a single gene (pedigrees 71, 239, ENG_AGZ), and one proband had variants in 2 genes that contributed to a blended phenotype (pedigree 269; variants in *ARMC4* and *ARMC9*). Additionally, 68/467 probands (14.6%) harbored 76 prioritized VUS SNV/indels among 52 genes; the VUS was compound heterozygous with a P/LP variant in pedigrees 193 and ENG_AZW. In pedigree ENG_COX, only a single allele was identified in a gene for which biallelic variants typically cause the phenotype. Some VUS have more supportive evidence, suggesting higher likelihood of their being substantiated over time (Table 1).

### SV analyses and classification with ACMG/ClinGen criteria

In 22 pedigrees, exome/genome analyses detected 21 rare candidate SVs that were predicted to perturb protein-coding sequences (Fig. 3E, Supplementary Results, Supplementary Table 10). We prioritized 5 SVs for ACMG/ClinGen classification^16^; these encompassed gene(s) associated with phenotypes consistent with the probands’. Three deletions among 4 probands were classified as pathogenic: an *HDAC8* deletion in pedigree 13 with syndromic sporadic ptosis, whose phenotype was consistent with *HDAC8*-associated conditions^37^; a deletion including *GCH1* in syndromic familial ptosis pedigree ENG_BS, which explained their ptosis and DOPA-related dystonia^38^; and a chr10q26 deletion in syndromic sporadic DRS pedigrees 233 and 131, which we reported recently (https://doi.org/10.1101/2023.12.22.23300468). The remaining two deletions scored as ACMG/ClinGen-VUS; they likely explained the clinical phenotypes but did not meet pathogenic classification thresholds. These included a partial deletion of *MACF1* in syndromic sporadic CN6-palsy pedigree 98, which we reported during the course of this study,^20^ and a partial deletion of *SALL4* in syndromic sporadic DRS pedigree ENG_DQ, whose phenotype was consistent with *SALL4-*associated conditions.^1^ See Table 1, Supplementary Table 7.

### Categorization of genes with causal and candidate SNVs/indels or SVs

The genes that harbored causal and candidate SNVs/indels or SVs (classified as P/LP or VUS, respectively) were identified through various combinations of analyses and fell into five distinct categories: 1) [oCCDD+,Syndrome+/-]: genes that were definitively associated with oCCDDs before this study and were genetically pre-screened in most probands. 2) [oCCDD(+),Syndrome+]: genes that had at least occasional prior oCCDD association but were typically part of specific monogenic syndromes and thus not pre-screened. 3) [oCCDD-,Syndrome+]: genes that fit the syndromic component of each proband’s phenotype but had no prior oCCDD association. 4) [oCCDD-,Syndrome-]: genes that had no reported association with either the oCCDD or non-CCDD phenotype of the probands who harbor them. 5) [Misdiagnoses]: genes associated with alternative non-neurogenic/ non-CCDD etiologies and represent misdiagnoses or oCCDD phenocopies. Genes harboring P/LP or VUS were predominantly in the [oCCDD(+),Syndrome+] category (Fig. 3F-G, Supplementary Results, Supplementary Table 7).

### Evaluation of classified or recurrent missense SNVs with AlphaMissense

To complement our genetic analyses, which depended on known biology and oCCDD etiologies, we evaluated ACMG/AMP-classified or recurrent missense variants with AlphaMissense (Fig. 3H, Supplementary Table 7).^12^ AlphaMissense labels predicted deleterious variants as “likely pathogenic” (LP), with the caveat that these pathogenicity assertions are not corroborated using non-computational evidence as for ACMG/AMP classifications. AlphaMissense scores 22.8 of 71 million potential proteome-wide missense variants as LP (32.1%).^12^ Of 41 ACMG/AMP-P/LP variants, 17 were missense and AlphaMissense scored 14 as LP (82.4%). Of 76 ACMG/AMP-VUS, 65 were missense and AlphaMissense scored 42 as LP (64.6%). Interestingly, among these 65 VUS, we highlighted 12 with especially strong genotype/phenotype correlations (Table 1); AlphaMissense scored 11 of these as LP (91.7%). Of the remaining 53, AlphaMissense scored only 31 as LP (58.5%). Finally, of 823 recurrent heterozygous missense variants among all 467 probands, 743 could be evaluated by AlphaMissense. Of these, only 83 scored as LP (11.2%); this count included a recurrent ACMG/AMP-pathogenic *KIF21A* p.(Arg954Trp) variant. AlphaMissense scores support our prioritized variants as deleterious, and also suggest that recurrent heterozygous variants may not be a large genetic contributor.

We next evaluated missense variants within the 5 gene categories above with AlphaMissense (Fig. 3H). In [oCCDD+,Syndrome+/-] genes – which had known prior oCCDD associations – 10/11 variants were scored as LP by AlphaMissense (90.9%); only 3 were ACMG/AMP-P/LP, suggesting that the remaining VUS are strong candidates for future study. AlphaMissense scored 1/11 variants as likely benign: a *TUBB3* ACMG/AMP-VUS p.(Cys124Ser), which we had ACMG/AMP-classified because of *TUBB3*-CFEOM associations but we had felt was less compelling a priori. [oCCDD(+),Syndrome+] contained 20/35 AlphaMissense-LP variants (57.1%), 6 of which were also ACMG/AMP-P/LP. Interestingly, two [oCCDD(+),Syndrome+] variants in *PTPN11* and *TRPV4* were ACMG/AMP-pathogenic but scored as ambiguous by AlphaMissense. In [oCCDD-,Syndrome+], 5/9 variants (55.6%) were AlphaMissense-LP, 3 of which were also ACMG/AMP-pathogenic, and a variant in *SCN1A* was pathogenic by ACMG/AMP but likely benign by AlphaMissense. Interestingly, [oCCDD-,Syndrome-], encompassing genes with no reported association with either the oCCDD or non-CCDD phenotypes, included 18/22 AlphaMissense-LP variants (81.8%); all were ACMG/AMP-VUS. Finally, in [Misdiagnoses] were 2/4 AlphaMissense-LP variants (50.0%), both of which were ACMG/AMP-P/LP. Cumulatively, these results suggest that AlphaMissense may aid in prioritizing ACMG/AMP-VUS.

### Characteristics of oCCDD probands with ACMG/AMP or ACMG/ClinGen P/LP variants

Collectively, SNV/indel and SV analyses identified ACMG/AMP/ClinGen-P/LP variant(s) in 43/467 pedigrees (9.2%; Fig. 3I, Supplementary Results, Supplementary Fig. 10, Supplementary Table 7). These variants explained the phenotype fully in 13 pedigrees and partially in 30 (Table 1). In the partially explained pedigrees, putative phenotype expansions were identified for oCCDDs (n=13), syndromic phenotypes (n=3), or both oCCDDs and syndromic phenotypes (n=14). In two cases, the P/LP allele was compound heterozygous with a VUS. Fourteen highly prioritized VUS were identified in 13 genes among 14 pedigrees (Table 1).

Moreover, we identified additional VUS in known and novel candidate genes which we believe merit future study (Supplementary Table 11).

P/LP variants were more common in probands with syndromic than isolated oCCDDs (38/197 versus 5/270; chi-square test of independence, Χ^2^=41.43, df=1, p=1.2e^-10^). The most common affected body systems were CNS (27/38, 71.1%), PNS/muscle/connective tissue (21/38, 55.3%), craniofacial (19/38, 50.0%), skeletal (non-scoliosis) (12/38, 31.6%), and skeletal (scoliosis) (12/38, 31.6%; Fig. 3I). Probands with P/LP variants had a mean of 3.67 affected body systems, and 25/38 syndromic probands (65.8%) had >2 body systems affected.

While P/LP variants were higher among familial than sporadic pedigrees (12/101, 11.9%; 31/366, 8.5%), most probands with P/LP variants had sporadic syndromic oCCDDs (29/43, 67.4%). Though bilateral oCCDDs were infrequent in our overall cohort, they were common among probands with P/LP variants (126/467 versus 19/43; chi-square test of independence, Χ^2^=5.73, df=1, p=0.017). Rates of P/LP variants varied among oCCDD subphenotypes (Fig. 3I).

## DISCUSSION

As the largest oCCDD cohort reported to date, our study contributes detailed clinical and MRI data to our public exome/genome datasets. Moreover, within the cohort we collate (1) manually curated genes that harbor variants and have putatively relevant animal models; (2) genes and pathways highlighted by GO analyses of DNVs; (3) prioritized pathogenicity-classified variants with comprehensive genotype/phenotype correlations; and (4) AlphaMissense scoring of highlighted missense variants. These resources should facilitate future oCCDD gene discovery.

Though most of our cohort was prescreened for variants in known oCCDD genes to enable novel gene discovery, 9.2% of our cohort had P/LP variants – many of which were not in our original list of oCCDD genes to pre-screen – and additional cases had strong VUS. Many pedigrees had diverse syndromic phenotypes and were sequenced to identify overlap between oCCDDs and other congenital defects; indeed, exomes/genomes identified diverse genes and pathways associated with many Mendelian conditions. These often had known roles in syndromic phenotypes but unknown or infrequent prior oCCDD connection ([oCCDD(+),Syndrome+], [oCCDD-,Syndrome+]). While some of these may explain only the syndromic phenotypes in the probands who harbor them, others likely represent true oCCDD phenotype expansions for known disorders. Examples included candidate variants in *MYH10* or in the TGF-beta pathway. MYH10 regulates actin organization, primary ciliary formation, and CN7 motor neuron migration.^25,39^ TGF-beta is expressed in motor neuron-adjacent mesenchyme^40^ and may act through a non cell-autonomous mechanism. We suggest additional studies of *MYH10* and TGF-beta genes in oCCDDs.

Our study has expanded the repertoire of variants in tubulin-encoding genes and identified multiple additional novel oCCDD candidate genes and variants, some of which had no prior human disease associations (e.g., *KLB, FGF21,* Supplementary Table 11). Future cross-phenotype examinations with other syndromic birth defect cohorts may solidify associations and identify additional shared etiologies, and ACMG/AMP/ClinGen and AlphaMissense classifications may prioritize the variants with highest potential.

Multiple factors likely contribute to the remainder of our cohort being unsolved. We used conservative classification criteria to mitigate overestimation of pathogenicity and only considered P/LP variants to fully or partially explain the phenotypes of the probands who harbored them. However, we identified many strong VUS candidates, some of which are particularly compelling and likely to become diagnostic over time (Table 1) or worthy of future novel candidate variant/gene research (Supplementary Table 11). Our cohort is phenotypically heterogeneous, suggesting that locus and allelic heterogeneity with frequent family-private variants could complicate identification of common genetic etiologies. This problem is not unique to oCCDDs, as exomes/genomes have exhausted large pedigrees and major genetic contributors to rare Mendelian conditions over time. (Fig. 3I). Additionally, since our study was restricted to coding variants, cases could be explained by other etiologies, including noncoding variants which remain challenging to interpret (https://doi.org/10.1101/2023.12.22.23300468). Finally, most probands had unilateral and sporadic oCCDDs, suggesting a lower likelihood of germline Mendelian genetic etiologies.

Limitations included bias toward genetically unsolved, syndromic, and sporadic cases; thus, our findings are not reflective of oCCDDs in the general population, many of which result from previously identified genetic etiologies. Moreover, because our cohort is predominantly of European descent, our study may miss other population-specific variants.

Our study expands the phenotypic spectrum of oCCDDs and elucidates missing genetic causes. This informs understanding of neurodevelopmental genetics and identifies novel genes and pathways to be prioritized in future studies.

## Data availability

Exome/genome sequencing data are accessible under dbGaP accession numbers phs001247.v1.p1 and phs001272.v2.p1. SNVs, indels, and SVs that were newly ACMG/AMP/ClinGen-classified by our study were submitted to ClinVar (Accession IDs: SUB14307097, SUB14310205, SUB14279226). A subset of variants were previously interpreted by the ACMG/AMP criteria by our team and submitted to ClinVar under separate accession IDs (pedigree ENG_1580, SCV001445961.1; pedigree 42, SCV001445941.1; pedigree ENG_CMO, SCV003761257.1; pedigree 71, SCV002507051.1 and SCV000693896.1; pedigree 144, SCV001430799.1; pedigree 38, SCV001449530.1). A variant in pedigree 13 was previously interpreted by the Undiagnosed Diseases Network for the same individual as we report in our cohort (SCV000837689.1).

## Acknowledgements

We thank our research participants and their families and Allison Rose, Kelsey McEntee, Victoria Timmel, Winnie Tung, and Narisu Narisu.

## Author contributions

Conceptualization: J.A.J., E.C.E.; Data curation: J.A.J., B.J.B., W.M.C., L.P., E.M.E., A.S.L., M.W.W., B.W., M.L.; Formal analysis: J.A.J., L.P., E.M.E., A.S.L., M.C.W., E.G., K.A.R., C.G., M.S.B., A.S.J., S.A.D.G., S.S., V.R.B, G.L., J.M.F., I.W., X.Z., H.B.; Funding acquisition: J.A.J., M.E.T., D.G.M., A.O’D.L., E.C.E.; Investigation: J.A.J., B.J.B., W.M.C., L.P., E.M.E., A.S.L., M.C.W., S.M., D.G.H., C.D.R., E.G., K.A.R., C.G., M.S.B., A.S.J., S.A.D.G., C.A., S.S., V.R.B., G.L., N.M., J.M.F., I.W., X.Z., B.M.P., M.M.W., S.B., P.M.M.R., L.B., E.E.F., C.R.Q., F.M.M., M.T., T.M., M.W.W., H.B., E.C.E.; Methodology: J.A.J., M.W.W., B.W., M.E.T., D.G.M., A.O’D.L., E.C.E.; Project administration: J.A.J., B.J.B., W.M.C., C.A., N.M., M.E.T., H.B., D.G.M., A.O’D.L., E.C.E.; Resources: M.E.T., D.G.M., A.O’D.L., E.C.E.; Software: J.M.F., I.W., X.Z., M.W.W., B.W., M.L., H.B.; Supervision: J.A.J., M.E.T., H.B., D.G.M., A.O’D.L., E.C.E.; Validation: J.A.J., W.M.C., E.C.E.; Visualization: J.A.J.; Writing-original draft: J.A.J., E.C.E.; Writing-review & editing: J.A.J., B.J.B., W.M.C., L.P., E.M.E., A.S.L., M.C.W., S.M., D.G.H., C.D.R., E.G., K.A.R., C.G., M.S.B., A.S.J., S.A.D.G., C.A., S.S., V.R.B, G.L., N.M., J.M.F., I.W., X.Z., B.M.P., M.M.W., S.B., P.M.M.R., L.B., E.E.F., C.R.Q., F.M.M., M.T., T.M., M.W.W., B.W., M.L., M.E.T., H.B., D.G.M., A.O’D.L., E.C.E.

## Ethics declaration

This study was conducted in accordance with the Declaration of Helsinki and approved by the Institutional Review Board of Boston Children’s Hospital (BCH), Boston, MA (protocol 05-03-036R). Data were collected in accordance with ethical guidelines of BCH. Written informed consent was obtained by qualified individuals for study participation. Participants’ genomic and clinical data remained linked but were deidentified.

## ADDITIONAL FILES

Supplementary Methods and Results (note: Supplementary Tables 2-4 and 11 are included here)

Supplementary Table 1. Demographics and phenotypes of probands and affected relatives in the sequenced cohort

Supplementary Table 5. Brain/orbital MRI findings

Supplementary Table 6. SNVs/indels identified in pedigrees with at least 3 members sequenced

Supplementary Table 7. Genetic findings in the cohort and genotype/phenotype correlations

Supplementary Table 8. Candidate genes with putatively relevant animal models

Supplementary Table 9. Gene ontology analysis of genes with de novo SNV/indels

Supplementary Table 10. Structural variants identified in the cohort

## Ocular CCDD Phenotyping Consortium

Hugo Abarca-Barriga, Christiane Al-Haddad, Jeffrey L. Berman, Erick D. Bothun, Jenina Capasso, Oscar Francisco Chacon-Camacho, Lan Chang, Stephen P. Christiansen, Maria Laura Ciccarelli, Monique Cordonnier, Gerald F. Cox, Cynthia J. Curry, Linda R. Dagi, Thomas Lee Dahm, Karen L. David, Bradley V. Davitt, Teresa De Berardinis, Joseph L. Demer, Julie Désir, Fabiana D’Esposito, Arlene V. Drack, Eric Eggenberger, James E. Elder, Alexandra T. Elliott, K. David Epley, Hagit Baris Feldman, Carlos R. Ferreira, Maree P. Flaherty, Anne B. Fulton, Christina Gerth-Kahlert, Irene Gottlob, Stephen Grill, Dorothy J. Halliday, Frank Hanisch, Eleanor Hay, Gena Heidary, Christopher Holder, Jonathan C. Horton, Alessandro Iannaccone, Sherwin J. Isenberg, Suzanne C. Johnston, Alon Kahana, James A. Katowitz, Melanie Kazlas, Natalie C. Kerr, Virginia Kimonis, Melissa W. Ko, Feray Koc, Dorte Ancher Larsen, Guillermo Lay-Son, Danielle M. Ledoux, Alex V. Levin, Richard L. Levy, Christopher J. Lyons, David A. Mackey, Adriano Magli, Iason S. Mantagos, Candice Marti, Isabelle Maystadt, Fiona McKenzie, Manoj P. Menezes, Claudia N. Mikail, David T. Miller, Kathryn Bisceglia Miller, Monte D. Mills, Kaori Miyana, H. U. Moller, Lisa Mullineaux, Julie K. Nishimura, A. Gwendolyn Noble, Pramod Kumar Pandey, Piero Pavone, Johann Penzien, Robert Petersen, James A. Phalen, Annapurna Poduri, Claudia R. Polo, Lev Prasov, Feliciano J. Ramos, Maria Ramos-Caceres, Richard M. Robb, Béatrice Rossillion, Mustafa Sahin, Harvey S. Singer, Lois E. H. Smith, Jeffrey A. Sorkin, Janet S. Soul, Sandra E. Staffieri, Heather J. Stalker, Steven F. Stasheff, Sonya Strassberg, Mitchell B. Strominger, Deepa Ajay Taranath, Ioan Talfryn Thomas, Elias I. Traboulsi, Maria Cristina Ugrin, Deborah K. VanderVeen, Andrea L. Vincent, Marlene C. Vogel G., Bettina Wabbels, Agnes M. F. Wong, C. Geoffrey Woods, Carolyn Wu, Edward Yang, Alison Yeung, Terri L. Young, Juan C. Zenteno, Alexandra A. Zubcov-Iwantscheff, Johan Zwaan

## SUPPLEMENTARY METHODS

### Cohort enrollment, data collection, and phenotyping

This study of 467 pedigrees was derived from a large cohort of individuals with oCCDDs and their relatives enrolled in our research study at Boston Children’s Hospital from August 1992 through June 2019. Individuals with the following oCCDD diagnoses were included: CFEOM, ptosis, MGJWS(+)ptosis, MGJWS(-)ptosis, INV-MGJWS(-)ptosis, CN4-palsy, Brown syndrome, DRS, CN6-palsy, HGP, or CCDD-NOS. Individuals with Moebius syndrome were not included in the present study. This study was conducted in accordance with the Declaration of Helsinki and approved by the Institutional Review Board of Boston Children’s Hospital (BCH), Boston, MA (protocol 05-03-036R). Data were collected in accordance with the ethical guidelines of BCH. Written informed consent was obtained by qualified individuals for study participation.

Participants’ genomic and clinical data remained linked but were deidentified.

Many but not all study participants (403/467 probands; 86.3%) were pre-screened for pathogenic variants in reported oCCDD disease genes (*KIF21A, PHOX2A, TUBB3, TUBB2B, CHN1, MAFB, SALL4, HOXA1*, *ROBO3*, and/or *ACKR3*) if their oCCDD phenotype described on initial enrollment was consistent with the phenotype attributed to the oCCDD gene.

Screening approaches varied based on available technologies at the time but included exome sequencing and Sanger sequencing validation, Sanger sequencing, ddPCR, Fluidigm, Haloplex, long-range PCR, qPCR, DHPLC, Surveyor, and SSCP. Pedigrees qualified for exome sequencing or genome sequencing (ES/GS) if they screened negative for common genetic etiologies, had sufficient DNA quality and quantity, and consented to broad genomic data sharing. Among these, pedigrees were prioritized if the oCCDD was familial, if the phenotype included syndromic features, and if the proband had been screened for variants in the common oCCDD genes.

Demographics including race, ethnicity, and biological sex assigned at birth were collected via survey and self-reported by study participants or their parents/legal guardians. These data were collected as required by the US National Institutes of Health and serve to contextualize the generalizability and limitations of our findings. Biological sex assigned at birth is reported at the individual and cohort levels, and race and ethnicity are reported at the cohort level only.

Self-reported categories of race included American Indian or Alaska Native, Asian, Black / African American, Native Hawaiian / Other Pacific Islander, More Than One Race, or White per the NIH Policy on Reporting Race and Ethnicity Data. Self-reported ethnicities included Hispanic or Latino, and Non-Hispanic or Latino.

Phenotypic data were obtained through retrospective review of clinical records, questionnaires, and clinical updates from research participants and their clinicians. Some probands self-acquired and shared ocular motility photographs using StrabisPIX, a HIPAA-compliant ophthalmologic application. When available, ocular motility data, photographs, and videos were reviewed retrospectively by a team of ophthalmologists, orthoptists, and neurologists (authors DGH, MCW, SM, ECE).

Affected individuals were assessed for presence or absence of phenotypes in 20 broad categories: restricted vertical eye movement, eyelid ptosis, restricted horizontal eye movement, synkinetic eye/facial movements, facial paralysis, hearing impairment, lower cranial nerve abnormalities (CN IX-XII), central nervous system (CNS) structural/functional malformation, malformations or dysfunction of additional peripheral nervous system (PNS)/muscle/connective tissue, craniofacial dysmorphisms, non-craniofacial dysmorphisms, skeletal abnormalities (non-scoliosis), skeletal abnormalities (scoliosis), pulmonary/lung/respiratory abnormalities, cardiovascular abnormalities, gastrointestinal (GI) abnormalities, renal/urinary/genital abnormalities, endocrine abnormalities, skin/hair/teeth/nail abnormalities, and other dysmorphisms/involvement. Data were not available for all categories for all participants.

When available, brain magnetic resonance images (MRIs) were obtained clinically and reviewed retrospectively by a pediatric neuroradiologist (author CR) for cranial nerve/ extraocular muscle findings and imaging quality, structural brain abnormalities, and abnormalities in other features including the orbits, globes, muscles of mastication, facial muscles, temporal bones, inner ear, internal auditory canals, skull, craniofacial shape, and spine. MRIs obtained by 3T imaging with thin slices (∼1 mm) were considered optimized for cranial nerve / extraocular muscle detection. When available, other imaging such as spinal MRIs or magnetic resonance angiography were reviewed for additional anomalies of the spine or vasculature, respectively. CN 1-12 were each read as normal, thin, absent/not visualized, or abnormal trajectory. Visible extraocular muscles were assessed for presence/ absence, size, and shape. Brain structures were assessed including cortex, corpus callosum, anterior commissure, basal ganglia, hippocampi, olfactory system, cerebral ventricles, white matter, myelination, cerebellar hemispheres, cerebellar vermis, midbrain, pons, and medulla.

In most pedigrees, the proband was designated as the individual in the pedigree who was first referred to our research team. In 6 pedigrees (ENG_CMV, ENG_CKA, ENG_VC, ENG_0686, ENG_1211, and ENG_2228), DNA from the true proband was not sequenced, and thus a secondary affected individual whose DNA was sequenced was designated as the proband.

Probands and pedigrees were assigned oCCDD diagnoses based on the oCCDD present in the proband. Mixed intrafamilial oCCDD presentations and oCCDD laterality for each proband and affected individual are noted (Supplementary Table 1).

Individuals were categorized as isolated if they had an oCCDD but no additional non-ocular congenital findings. Individuals were categorized as syndromic if they had convincing documentation of at least one major or two minor congenital anomalies in addition to their oCCDD and/or if clinical genetic evaluation included chromosomal/genetic analysis for syndromic features. Major and minor congenital anomalies were defined using the CDC Birth Defects Surveillance Toolkit.^1^ Scoliosis was counted as a major anomaly if it required medical intervention. Hearing impairment was counted as a major anomaly if attributable to non-conductive etiologies, including sensorineural deficits or congenital ear malformations. Syndromic findings that were explained by an alternative genetic diagnosis are denoted.

Individuals who lacked a family history of oCCDDs or related findings were categorized as sporadic. Individuals with a family history of oCCDDs were designated as familial. Pedigree structures were assessed to determine the most likely mode(s) of inheritance.

If both the proband and one or more relatives had a syndromic oCCDD, the pedigree was designated as “syndromic familial” and both the oCCDD and syndromic features were documented as the heritable components in the pedigrees. If, however, the proband had a syndromic oCCDD and had relatives with the syndromic features in the absence of the oCCDD, the pedigree was designated as “syndromic familial” and the non-CCDD phenotype was documented as the heritable component in the pedigree (Supplementary Table 1). In such pedigrees, only the proband was designated as “affected” for counting purposes, since only the proband had an oCCDD. Pedigrees were designated as “isolated” if syndromic feature(s) were absent in the proband and present in family members who did not have oCCDDs.

For pedigrees with highlighted genetic findings, individual clinical features were reported using Human Phenotype Ontology (HPO) terms.^2^ Certain findings were noted clinically but not included as separate HPO terms if these findings are frequently encompassed by the overarching oCCDD diagnosis (e.g., esotropia, exotropia, amblyopia, and ptosis frequently accompany CFEOM). Highlighted cases were assessed for correlative phenotypic data to obtain supportive evidence for or against candidate gene/variant causality.

### Co-occurring defects analysis (CODA) of oCCDDs and syndromic phenotypes

Co-occurring defects analysis (CODA) was used to determine whether oCCDDs and syndromic phenotypes co-occurred more frequently than by chance.^3^ Separate analyses of syndromic phenotypes were performed for each of the most common oCCDD subdiagnoses (DRS, CFEOM, and congenital ptosis). CODA examines the observed number of cases for all combinations of two through five co-occurring phenotypes and determines whether each combination occurs more frequently than if the phenotypes were independent. The method adjusts for the tendency of birth defects to co-occur with other birth defects and produces an adjusted observed/expected ratio (OEadj). OEadj values >1, which indicate enrichment of the co-occurrence of the given phenotypes, were retained if present in at least 3 probands for DRS (n=79 total syndromic probands), at least 3 probands for CFEOM (n=50 total syndromic probands), or at least 2 probands for congenital ptosis (n=26 total syndromic probands).

Partially redundant phenotype combinations were removed. Syndromic phenotypes used for CODA included 15 of the 20 syndromic categories defined in the “Cohort enrollment, data collection, and phenotyping” section above. The miscellaneous “other findings’’ category of phenotypes was excluded due to large amounts of missing data. Four of the 20 syndromic categories were also excluded since they define specific oCCDD subcategories and thus provide less informative associations: restricted vertical motility, restricted horizontal motility, synkinesis, and ptosis.

### DNA sample collection and sequencing

Of 1310 genetically unsolved pedigrees in our cohort, 467 underwent exome/genome sequencing and phenotypic analyses for this study (550 affected and 1108 total individuals). DNA was extracted from blood or saliva of oCCDD probands and their relatives using Puregene manual blood extraction or prepIT.L2P saliva DNA extraction solution (DNA Genotek, Ottawa, ON, Canada). Each pedigree underwent exome or genome sequencing. Exome sequencing was conducted through the Center for Mendelian Genomics (CMG) at the Eli and Edythe L. Broad Institute of MIT and Harvard (Broad CMG; Cambridge, MA, USA). Genome sequencing was conducted through the Broad CMG or through the NIH Gabriella Miller Kids First (GMKF) Pediatric Research Program at the Baylor College of Medicine Human Genome Sequencing Center (Houston, TX) and then reprocessed by the Broad CMG. We refer to these sequencing datasets as CMG_ES, CMG_GS, and GMKF_GS, respectively.

The three datasets were generated at different times through separate collaborative initiatives. As such, each had unique selection criteria that determined whether samples underwent exome or genome sequencing. The first dataset sequenced was GMKF_GS, which prioritized pedigrees for genome sequencing for which adequate DNA and data sharing consents were available for at least 3 pedigree members. In addition, pedigrees with syndromic oCCDDs were prioritized to search for overlap with other developmental phenotypes. The second genomed dataset was CMG_GS, which permitted genome sequencing of a smaller number of samples and was aimed at identifying additional alleles in candidate genes identified in familial pedigrees sequenced through GMKF_GS. Accordingly, this dataset was predominantly composed of singletons with oCCDDs and a few additional larger pedigrees with adequate DNA and consents. The final dataset generated was CMG_ES, which permitted exome sequencing of a larger number of samples with variable phenotypes and pedigree structures that had adequate DNA and consents. Consequently, this cohort included heterogeneous samples that had not been sequenced in either of the aforementioned genome sequencing datasets.

DNA libraries were generated for CMG_ES with a 38-Mb target Illumina exome capture and sequenced with 150 bp paired-end reads to cover >80% of targets at 20x, with an average target coverage >55x. DNA libraries were prepared for GMKF_GS with the KAPA Hyper PCR-free library prep kit (KAPA Biosystems Inc., Wilmington, MA) and sequenced on the Illumina HiSeq X to 30X average coverage using 150 bp paired-end reads. PCR-free DNA libraries were made for CMG_GS using Illumina HiSeq X Ten v2 and sequenced with an average target coverage >30x. Quality assurance checks were performed to confirm sample identity and parentage when appropriate. All datasets were then processed at the Broad using a Picard-based pipeline (http://broadinstitute.github.io/picard/) integrating base quality score recalibration and local realignment around indels. Reads were mapped to the human reference genome (GRCh38)^4^ using BWA.^5^ Single nucleotide variants (SNVs) and small insertions and deletions <50bp (indels) were called jointly among >10,000 exomes or genomes assembled by the Broad using Genome Analysis Toolkit (GATK) HaplotypeCaller v4.0 (GMKF_GS and CMG_GS) or v3.4 (CMG_ES).^6,7^ SNVs and indels were filtered using default GATK Variant Quality Score Recalibration parameters. Variants were annotated with Ensembl Variant Effect Predictor^8^ and analyzed with seqr (https://seqr.broadinstitute.org/).^9^

### Genetic imputation of global ancestry

Global genetic ancestry was inferred from exome/genome sequences using principal component analysis as previously described.^10^ Individuals were classified into the following genetically imputed ancestral groups: African/ African American, Amish, Ashkenazi Jewish, East Asian, European (Finnish), European (non-Finnish), Latino/Admixed American, Remaining Individuals, or South Asian. These ancestral genetic groups were reported at an aggregate level for the cohort.

### SNV/indel filtering and prioritization

For each pedigree, SNVs and indels were analyzed according to the most probable mode(s) of inheritance based on pedigree structure using seqr (https://seqr.broadinstitute.org/).^9^ This included custom analyses to account for observed incomplete penetrance, when present. Using 1000 Genomes Project Phase 3,^11^ ExAC v0.3,^12^ gnomAD v2.0.2,^10^ and TOPMed Freeze 5^13^ reference datasets, we identified homozygous or compound heterozygous variants with allele frequencies (AF) <0.01 under an autosomal recessive (AR) model or heterozygous variants with AF<0.001 under autosomal dominant (AD), *de novo*, or X-linked recessive (XLR) models.

Variants that were homozygous in reference population(s) were excluded. Heterozygous variants present in >5 individuals and all variants with AF>0.01 in an internal database of rare disease samples from the Broad CMG were excluded. Variants that passed quality control filters and had allele balances >25 and genotype qualities >20 were selected. Indels and missense, nonsense, or essential (+/-2 bp) splice site-altering variants were prioritized in all annotated protein-coding genes. Broader noncoding variant analyses were conducted and reported separately (https://doi.org/10.1101/2023.12.22.23300468). Variant annotation was confirmed in June of 2023 with VariantValidator.^14^ MANE Select transcripts were prioritized. Lists of all variants meeting these parameters in singletons were generated, but singleton sequences were assessed only for variants in known oCCDD genes or strong candidate genes and for variants annotated as pathogenic/ likely pathogenic in ClinVar in additional genes.

### Identification of recurrently mutated genes

Genes that had SNVs/indels meeting the parameters defined in “SNV/indel filtering and prioritization” that were mutated in more than one pedigree were defined as “recurrently mutated genes.” Recurrently mutated genes were queried in all 467 pedigrees regardless of specific oCCDD diagnosis.

### Biological prioritization of SNVs/indels

Further qualitative prioritization was performed of candidate variants based on their CADD v1.6 PHRED scores.^15^ Candidate genes were further prioritized based on their LOEUF^10^ and missense z-scores;^16^ functions annotated in the primary literature, and prior reported associations with human phenotypes in Online Mendelian Inheritance in Man (OMIM, www.omim.org),^17^ the Human Gene Mutation Database (HGMD, www.hgmd.cf.ac.uk; license #7597755),^18^ and ClinVar^19^ databases and in the primary literature. Select variants were prioritized based on: prior reporting in unrelated individuals in the literature or in ClinVar; consistency of the phenotypes in our proband(s) relative to previously reported probands with variants in the same gene; or of identification of multiple rare, putatively damaging alleles in unrelated individuals with similar phenotypes in our cohort.

### DenovolyzeR analysis of de novo variants

Statistical analysis of SNV/indel de novo variants (DNVs) was performed with DenovolyzeR.^20^ 203 pedigrees were amenable to DNV analysis and comprised a minimum structure of 2 unaffected parents and 1 affected child (129 from GMKF_GS; 6 from CMG_GS; 68 from CMG_ES). If additional relatives were also sequenced outside of the nuclear trio, these members’ sequences were omitted. Pedigrees with 2 affected siblings of two unaffected parents were excluded from the analysis, thus omitting pedigrees with putative germline mosaicism. In total, 197 trios, 4 quads, and 2 pedigrees with other pedigree structures (>4 sequenced individuals) were used. The individuals whose sequences were used for these analyses are specified (Supplementary Table 1, Column I). oCCDDs in these pedigrees were variable (51 isolated DRS, 36 syndromic DRS, 12 isolated CFEOM, 28 syndromic CFEOM, 9 isolated congenital ptosis, 11 syndromic congenital ptosis, 13 isolated MGJWS(+)ptosis, 5 syndromic MGJWS(+)ptosis, 3 isolated MGJWS(-)ptosis, 2 syndromic MGJWS(-)ptosis, 1 isolated INV-MGJWS(-)ptosis, 11 isolated CN4-palsy, 10 syndromic CN4-palsy, 2 isolated Brown syndrome, 2 syndromic Brown syndrome, 3 syndromic horizontal gaze palsy, 2 syndromic CN6-palsy, 2 syndromic CCDD-NOS).

Heterozygous autosomal missense, nonsense, canonical splice site, synonymous, and frameshifting SNVs/indels were selected that occurred de novo in the proband and had AF<0.001 in the 1000 Genomes Project Phase 3,^11^ ExAC v0.3,^12^ gnomAD v2.0.2,^10^ and TOPMed Freeze 5^13^ databases and <0.01 in an internal database of rare disease samples. Variants were retained that passed filters and had genotype quality >30, allele balance >25, read depth >10, and alternate allele count ≥5. Three pedigrees (67, 252, and 14) had DNV counts > 3 SD from the mean and were considered outliers and excluded from further analysis.

Variants were aggregated into classes based on predicted loss-of-function (pLOF; nonsense, splicing, frameshift) or missense consequences. If multiple DNVs of any class were represented more than once in a single proband (e.g., 2 missense variants in the same gene in one person), only one variant was counted for downstream analyses. Through denovolyzeR, we used the functions denovolyzeByClass to assess overall enrichment of various classes of DNVs, denovolyzeMultiHits to assess enrichment of recurrently mutated genes with DNVs in any class, and denovolyzeByGene to assess whether enrichment within any class was attributable to recurrent DNVs in any individual genes as opposed to collective DNVs across multiple genes.

### Gene ontology analysis of DNVs

DNVs were obtained for gene ontology (GO) analysis as described in the denovolyzeR methods above, and synonymous variants were removed. Functional enrichment analysis was performed using g:Profiler g:GOSt (version e110_eg57_p18_4b54a898) with g:SCS multiple testing correction and a significance threshold of 0.05.^21^ P-values are provided as adjusted p-values (p_adj_) obtained after multiple test correction.

### Coding structural variant (SV) analysis

In samples with genome sequencing, structural variants (SVs) that perturb the coding sequence were identified, jointly genotyped, and annotated using the ensemble SV discovery tool GATK-SV (https://github.com/broadinstitute/gatk-sv).^22^ Each cohort was processed with samples from pedigrees with unrelated rare disorders to serve as controls for improved filtering. SVs in each oCCDD pedigree were analyzed according to the most probable mode(s) of inheritance based on pedigree structure. *De novo* SVs were detected using a published pipeline that applies a series of post-hoc filters to a GATK-SV VCF.^23^ Rare inherited SVs were identified by filtering on an AF<0.005 based on unaffected individuals in our internal cohort. In addition, the gnomAD SVs v2.1 database was used for AF filtering using the same cutoffs defined for SNVs/indels. Partial or full deletions of ≥1 exon and duplication or inversion events with exonic breakpoint(s) were prioritized for pathogenicity review. SVs were visually inspected with IGV^24^ and the RdTest module in GATK-SV for validation, and passing variants were prioritized. Genes within each SV interval were evaluated for predicted haploinsufficiency or triplosensitivity^25^ and biological relevance, recurrent disruption of the same gene(s) among multiple pedigrees, and putatively relevant animal models, as defined in “animal model analyses” described below.

In samples with exome sequencing, SVs consisting of rare coding deletions and duplications were delineated with the recently published GATK-gCNV algorithm.^26^ GATK-gCNV is a read-depth based method, specifically tailored to detect rare coding copy number variants (CNVs) from ES data with excellent sensitivity and false positive identification characteristics. In brief, we batched samples to be processed across different technical cohorts based on technical sequencing fluctuations such as those arising from a difference in sequencing center or in exome enrichment kits. Each of these technical cohorts was then relayed into the GATK-gCNV pipeline, which automatically normalized the read-depth signal onto the copy number scale and output highly sensitive candidate CNVs. A series of deeply benchmarked quality control filtering metrics, as outlined in our recent publication,^27^ was applied to prioritize CNVs. *De novo* status of these identified CNVs was determined in settings where we had complete trios by examining whether either of the parents harbored evidence of a matching CNV as the proband. Likewise, as with the samples with genome sequencing, genes within the identified CNVs were evaluated for predicted biological relevance, recurrent disruption of the same gene(s) among multiple pedigrees, and putatively relevant animal models, as defined in “animal model analyses” below.

### Animal model analyses

Using the thresholds for allele frequency, variant annotations, and quality control defined above in the “SNV/indel filtering and prioritization” and “Coding structural variant (SV) analysis” sections, recurrently mutated genes as well as strong candidate genes meeting criteria defined in “Biological prioritization of SNVs/indels” were annotated for putatively relevant animal model phenotypes in the Monarch database (http://monarchinitiative.org).^28^ Putatively relevant phenotypic terms were extremely diverse; examples included phenotypes in cranial nerves, motor neurons, other neurodevelopmental phenotypes, abnormalities of the orbital region, putatively relevant biological processes (e.g. involvement in axon growth/ guidance), and other syndromic features that are seen in some oCCDD cases (e.g. limb anomalies). We performed this analysis while bearing in mind the caveat that most animal models harbor loss-of-function alleles, while others have specific resulting in gain-of-function or altered function alleles, and these allelic consequences may or may not be consistent with the consequences of the variant(s) in our sequenced human cohort.

Following the identification of all genes harboring candidate variants that had compelling animal model data, we next evaluated each for full-gene and local missense constraint, predicted variant pathogenicity, population frequency, protein domain localization, relative location of pedigrees’ variants, and biological function. Human probands with variants in the same gene were evaluated for phenotypic consistency with one another and with their animal model orthologs. Genes/variants that were most compelling based on these collective criteria were highlighted.

### Genotype/phenotype correlations

For pedigrees with highlighted genetic findings obtained through analyses of genotype/phenotypes, animal models, and/or DNVs, clinical features were reported using Human Phenotype Ontology (HPO).^2^ Highlighted cases were assessed for correlative phenotypic data to obtain supportive evidence for or against candidate gene/variant causality.

### Interpretation of variant pathogenicity and submission to ClinVar

Through qualitative manual review of genes obtained from analysis of genotype/phenotypes, DNVs, and animal models, select variants were chosen for formal interpretation. SNVs and indels were interpreted in May of 2023 using recommendations from the American College of Genetics and Genomics and Association for Molecular Pathology (ACMG/AMP)^29^ in accordance with the standard operating procedure defined by the Clinical Genome Resource (ClinGen) Variant Curation Committee (ClinGen General Sequence Variant Curation Process Version 1.0; https://www.clinicalgenome.org/site/assets/files/3677/clingen_variant-curation_sopv1.pdf), using REVEL as the computational predictor for missense variation.^30^ SVs were interpreted in May of 2023 using joint recommendations from ACMG and the Clinical Genome Resource (ClinGen).^31^ The number of genes within each SV interval was annotated using OMIM. Variants were classified as pathogenic (P), likely pathogenic (LP), or variants of uncertain significance (VUS). A subset of variants were previously interpreted by the ACMG/AMP criteria by our team and submitted to ClinVar under separate accession IDs (Data Availability). Variant annotation was standardized using VariantValidator.^14^

### Variant categorization

The genes that harbored ACMG/AMP/ClinGen-classified SNVs/indels or SVs were grouped into five categories as detailed in main text and Figure 3:

1. [oCCDD+,Syndrome+/-]: genes that were definitively associated with oCCDDs before this study and were genetically pre-screened in most probands.
2. [oCCDD(+),Syndrome+]: genes that had at least occasional prior oCCDD association but were typically part of specific monogenic syndromes and thus not pre-screened.
3. [oCCDD-,Syndrome+]: genes that fit the syndromic component of each proband’s phenotype but that, to our knowledge, have no prior oCCDD association.
4. [oCCDD-,Syndrome-]: genes that, to our knowledge, had no reported association with either the oCCDD or non-CCDD phenotype of the probands who harbor them.
5. [Misdiagnoses]: genes associated with alternative non-neurogenic/ non-CCDD etiologies and represent misdiagnoses or oCCDD phenocopies.

### Identification of recurrent missense variants

Using the thresholds for allele frequency, variant annotations, and quality control defined above in the “SNV/indel filtering and prioritization” section, recurrent heterozygous variants were identified among DNVs in sporadic cases, autosomal dominant variants in familial cases, and heterozygous variants in duos/singletons. Variants were considered recurrent if mutated among >1 pedigree with any oCCDD subdiagnosis, not just among pedigrees with the same oCCDD. Recurrent missense variants were not prioritized for ACMG/AMP classification, but were scored by AlphaMissense, as defined in the following section.

### Evaluation of recurrent and prioritized missense variants with AlphaMissense

We scored both recurrent missense variants and ACMG/AMP-classified missense variants using AlphaMissense.^32^ The latter included all heterozygous, homozygous, or compound heterozygous missense variants that were represented in AlphaMissense and classified as pathogenic, likely pathogenic, or VUS by ACMG/AMP criteria (Supplementary Table 7).

ACMG/AMP/ClinGen-VUS that we felt were most compelling and featured in Table 1 were distinguished from additional VUS that currently have lower levels of evidence.

AlphaMissense predictions were based on default score cutoffs designed to reach 90% precision on ClinVar variants. Variants evaluated by AlphaMissense were concurrently stratified into the five categories described above to assess breakdown of AlphaMissense scores within each category. Additionally, AlphaMissense scores were compared to ACMG/AMP classifications.

### Figure generation

Components of figures were generated in R, and main and supplementary figures were assembled with BioRender.com using an academic license provided to Boston Children’s Hospital (figure license numbers: KM26LVQ8FB, QW26LVQ72D, FK26LVQ58S, YX26JQRKIF, CM26JQRIAP, AI26JQUI4A, JC26JQWDZS, VH26JQW1Z0, UZ26JQWNGK, OQ26KR14PB, BU26JQX3RC).

## SUPPLEMENTARY RESULTS

### oCCDD Phenotype Summaries in Sequenced Probands

Multiple oCCDDs were occasionally seen within a single pedigree (n=7, 1.5%). In 4 syndromic DRS pedigrees (0.9%), the syndromic non-CCDD component of the phenotype was inherited, but the oCCDD was present only in 1 affected individual in the pedigree (Supplementary Table 1). By subdiagnoses, oCCDDs were unilateral in 62.6% of DRS, 38.8% of CFEOM, 73.3% of congenital ptosis, 73.6% of MGJWS, 67.9% of CN4-palsy, 69.2% of Brown syndrome, 0% of horizontal gaze palsy, 40% of CN6-palsy, and 44.4% of CCDD-NOS probands (Fig. 2B, Supplementary Fig. 7, Supplementary Tables 1-2). In total, 284 of 467 probands (60.8%) had some form of synkinesis (Fig. 2C). By definition, this includes all probands with DRS and MGJWS. Synkinesis was also common in CCDD-NOS (88.9%) and CFEOM (28.8%), and less frequent in Brown syndrome (7.7%) and congenital ptosis without MGJWS (1.3%). Synkinesis was not documented in probands with CN4-palsy, horizontal gaze palsy, or CN6-palsy (Supplementary Fig. 8, Supplementary Tables 1-2). Synkinetic patterns varied with diagnosis and among probands (Supplementary Table 1).

### Syndromic/ Non-CCDD Phenotype Summaries

Among the 197 probands with a syndromic oCCDD, their syndromic findings were categorized as: CNS (55%), PNS/muscle/connective tissue (37%), craniofacial (37%), skeletal (24%), gastrointestinal (23%), cardiovascular (22%), skin/hair/teeth/nails (22%), other dysmorphisms (19%), renal/urinary/genital (17%), hearing (13%), endocrine (13%), scoliosis (12%), pulmonary/lung/respiratory (12%), facial paralysis (8%), and lower CN IX-XII (5%); the distribution of these syndromic findings were variable among oCCDD subdiagnoses (Fig. 2D, Supplementary Tables 1 and 3).

CODA analysis of co-occurring phenotypes among syndromic probands with DRS, CFEOM, or congenital ptosis yielded 156 non-redundant phenotype combinations with OEadj>1 that were present in at least 3 probands for DRS or CFEOM or 2 probands for congenital ptosis (n=76, 58, and 22 for DRS, CFEOM, and congenital ptosis, respectively; Supplementary Table 4). Among these significant results were patterns of co-occurring phenotypes underlying a few recognizable syndromes. For instance, individuals with DRS and 10q deletion syndrome had CNS structural/functional malformation, PNS/muscle/connective tissue, skeletal (scoliosis), and renal/urinary/genital involvement (OEadj=6.04); individuals with Duane radial-ray syndrome had skeletal and renal involvement (OEadj=1.71); and individuals with CFEOM and ciliopathies had CNS structural/functional malformation, PNS/muscle/connective tissue, craniofacial, skeletal (non-scoliosis), and endocrine involvement (OEadj=9.05). While CODA analysis highlighted that many probands shared groupings of affected systems, their specific underlying phenotypes or genetic diagnoses often differed, so this analysis did not identify additional novel syndromes.

We suggest that similar future analyses include more specific endophenotypes rather than broad systemic groupings to facilitate the identification of additional syndromes or to generate more informative pedigree groupings for analysis of shared genetic etiologies of syndromic phenotypes.

### Brain MRI Findings in Sequenced Probands

To the extent feasible, MRI scans were assessed for cranial nerve and/or extraocular muscle anomalies, additional brain anomalies, and other non-brain anomalies (Supplementary Table 5, Fig. 2E). In total, 81 probands (83 affected individuals) had brain and/or orbital MRIs, of which scans from 47 probands (49 affected individuals) were available for review. Of these 47 probands, 25 had CFEOM (53.2%) and 11 had DRS (23.4%), while 4 had congenital ptosis, 3 had CN6-palsy, 3 had CCDD-NOS, and one had Brown syndrome (Supplementary Table 5, Fig. 2E). Thirty-one scans permitted interpretation of cranial nerve and/or extraocular muscle anatomy, of which 13 were optimized for cranial nerve/ extraocular muscle detection (8 CFEOM, 4 DRS, and 1 CCDD-NOS).

#### Findings in internally reviewed MRIs

Six of eight CFEOM probands with optimized MRIs had detectable thinning or absence of CN3; in five, CN3-innervated extraocular muscle(s) could also be assessed and were small. Exceptions included two CFEOM probands who, contrary to expectation, had normal-appearing CN3. One of these had normal CN3 but small CN3-innervated extraocular muscles (pedigree 41), suggesting possible limitations in detecting cranial nerve misrouting, defasciculation, or thinning. The second CFEOM proband with normal-appearing CN3 had orbital bands tethering the extraocular muscles (pedigree 99), suggesting a non-CCDD etiology as the cause of the clinical oCCDD diagnosis. All four DRS probands with optimized MRIs had thin/ absent CN6; while two had thin lateral rectus muscles, the lateral rectus muscles were normal-appearing in two, consistent with maintenance of the muscle secondary to aberrant innervation by CN3 in DRS (Supplementary Table 5).

Among probands with optimized MRIs, 9/13 (69.2%) had consistent laterality between their clinically detected oCCDD versus their MRI-derived cranial nerve/ extraocular muscle abnormalities. Inconsistencies were detected in 2 CFEOM probands and included clinically bilateral oCCDDs but unilateral MRI findings (pedigrees ENG_ASV and 260), again suggesting limitations in resolving subtle cranial nerve thinning. Interestingly, the only pedigree with MRI for >1 sequenced individual (pedigree 260 with CFEOM) had a mixed intrafamilial presentation with variable brain and other findings on MRI among different members (Supplementary Table 5).

Scans revealed diverse combinations of additional structural brain anomalies in 34/47 probands (72.3%). Commonly affected structures across all oCCDDs included the corpus callosum (17/34, 50.0%), cerebral ventricles (14, 41.2%), cerebellum (14, 41.2%), cerebral cortex (11, 32.4%), anterior commissure (11, 32.4%), and hippocampus (10, 29.4%) (Fig. 2E; Supplementary Table 5). Cerebral cortex, pons, and olfactory system were also more frequently affected in DRS than in CFEOM, while anterior commissure and midbrain were more frequently affected in CFEOM than in DRS, but these associations were not statistically significant (chi-square test of independence, Fig. 2E). While some of these trends were expected (e.g. increased involvement of anterior commissure and midbrain in CFEOM), others were not (e.g. increased olfactory system involvement in DRS). This may be because imaged individuals often had atypical syndromic presentations, but may be in part because previous studies have not systematically ascertained these structures in all oCCDD subgroups.

Imaging revealed non-brain anomalies in 29/47 probands (61.7%). Common anomalies across all oCCDDs were in skull shape (9/29 31.0%), inner ear (7, 24.1%), vasculature (7, 24.1%), spine (6, 20.7%), internal auditory canals (4, 13.8%), and craniofacial structures (4, 13.8%) (Fig. 2E; Supplementary Table 5). While spine anomalies were more commonly observed in DRS than in CFEOM (3/7 versus 1/16; chi-square test of independence, Χ^2^=4.54, df=1, p=0.033), this may have been due in part to targeted imaging of the spine in these individuals. Craniofacial and skull shape anomalies and microcephaly were more common in CFEOM, while inner ear anomalies were more common in DRS, but these associations were not statistically significant on evaluation by the chi-square test of independence.

#### Findings in externally reviewed MRIs

We received written reports but were unable to obtain MRIs for review from 34 probands. Seven were reported to have cranial nerve/ extraocular muscle findings (cranial nerve abnormalities in 3; extraocular muscle abnormalities in 5). Of these, imaging was reported to be optimized for cranial nerve/ extraocular muscle detection in 2, both of which showed consistency in laterality of the clinically detected oCCDD and the cranial nerve/ extraocular muscle findings. Among externally reported MRIs, 10 had structural brain abnormalities. Common findings were in the corpus callosum (3/10, 30.0%), white matter volume (30.0%), cerebral cortex (20.0%), cerebellum (20.0%), pons (20.0%), and medulla (20.0%) (Supplementary Table 5).

### Pre-existing genetic diagnoses

Clinical genetic findings identified before exome/genome sequencing explained the syndromic non-CCDD phenotypes in two individuals. These were proband ENG_AKL, who had congenital ptosis and mosaic Turner syndrome, and affected individual 178_04, who was not the proband of their pedigree but had DRS and Klinefelter syndrome (Supplementary Table 1).

### Analysis of genes from SNV/indel analyses and their animal model phenotypes

The following 2000 genes had putatively relevant animal models in the Monarch database^28^ under each mode of inheritance: XLR (6 genes), AR (57 genes), and AD (1973 genes); 36 genes were represented in both the AD and AR categories. Some model phenotypes were in generic neurodevelopmental processes, while others were more specifically oCCDD-related (detailed phenotypes provided in Supplementary Table 8).

The variants/genes in each pedigree that had compelling animal model data were then evaluated for full-gene and local missense constraint, predicted variant pathogenicity, population frequency, protein domain localization, relative location of pedigrees’ variants, and biological function. Human probands with variants in the same gene were evaluated for phenotypic consistency with one another and with their animal model orthologs. This resulted in the prioritization of 95 variants in 59 genes among 89 pedigrees. Among these were candidate variants in genes without known human oCCDD involvement. The logic for prioritizing *NES, CUX1, GNAS, FER, ACTR1B, OLIG2,* and *SEMA3F* as putative novel candidate genes is provided below, while *KIF5C* is provided in the main text.

The intermediate filament protein-encoding gene *NES* harbored an AD variant in isolated familial CFEOM pedigree 251 that was rare and predicted damaging (c.23A>T, p.(Glu8Val), NM_006617.2; Supplementary Table 7). *NES* orthologous zebrafish mutants have multiple phenotypes, including small, disorganized, and apoptotic midbrain; multiple abnormal cranial nerves including CN3 and CN4; decreased neuronal precursors; and abnormal neuron differentiation (Supplementary Table 8).^33^ *NES* is also highly expressed in adult human extraocular muscles, suggesting that alternative non-CCDD etiologies could also be involved. However, this gene has a compelling variant in just one pedigree in our cohort.^34^

*CUX1*, encoding a transcription factor involved in neuronal differentiation, is mutated in 3 DRS pedigrees: isolated sporadic DRS singletons ENG_PQ (c.724A>G, p.(Met242Val)) and ENG_GH (c.3853A>G, p.(Ile1285Val)) and isolated familial DRS trio 230 (c.3793G>A, p.(Glu1265Lys), NM_001202543.2) (Supplementary Table 7). All three variants are rare or absent from population databases and have moderate to damaging predictions. Residues 1265 and 1285 are in the protein homedomain, whereas residue 242 is not in an annotated domain. *CUX1* is highly missense-constrained within humans (missense z=3.749).^16^ Notably, CUX1 binds to known DRS-associated protein CHN1,^35^ and a mutant *CUX1* fly ortholog has abnormal neuroanatomy and neurophysiology (Supplementary Table 8). Largely LOF DNVs in *CUX1* have been reported in global developmental delays with or without intellectual disabilities (MIM116896), a phenotype which was not documented in any of our three pedigrees.

*GNAS,* encoding a G-protein whose signaling modulates hormones and neurotransmitters, is mutated in 4 DRS singletons: isolated sporadic DRS probands ENG_UE (c.713G>A, p.(Gly238Glu), NM_016592.5), ENG_JU (c.1591C>T, p.(Pro531Ser), NM_080425.4), and ENG_AAJ (c.1717G>C, p.(Asp573His), NM_080425.4) and syndromic sporadic DRS proband ENG_KS (c.304G>C, p.(Glu102Gln), NM_016592.5) (Supplementary Table 7). *GNAS* is missense-constrained within humans (missense z= z-score: 2.655),^16^ and the variants are rare and have moderate to damaging predictions. Interestingly, many of the syndromic features of proband ENG_KS have been previously associated with *GNAS* variants^36^ (Supplementary Table 7), but DRS has not. Orthologous worm mutants have microtubule cytoskeleton abnormalities, and fly mutants have abnormal neurophysiology and smell perception (Supplementary Table 8).

*FER* is mutated in two singletons with isolated sporadic DRS (*FER*: ENG_1616, c.1883C>T, p.(Thr628Ile); ENG_1637, c.1887A>C, p.(Gln629His), NM_001308028.2; Supplementary Table 7). Both variants are absent from population databases and have moderate to damaging predictions. Interestingly, *FER* encodes a protein tyrosine kinase that regulates diverse processes including synaptic vesicle trafficking, actin cytoskeleton regulation, and microtubule assembly, and the *FER* variant residues in ENG_1616 and ENG_1637 are directly adjacent to one another in the protein kinase domain. *FER* worm models have abnormalities of axon midline crossing and receptor-mediated endocytosis (Supplementary Table 8).

*ACTR1B* is mutated in two singletons with isolated sporadic DRS (ENG_CMJ, c.1006C>G, p.(Arg336Gly); ENG_BAE, c.633T>A, p.(Phe211Leu), NM_005735.4; Supplementary Table 7). Both variants are absent from population databases and have moderate to damaging predictions. *ACTR1B* encodes a protein involved in vesicle movement along the microtubule, and worm models have defective receptor-mediated endocytosis (Supplementary Table 8).

*OLIG2* encodes a transcription factor and is mutated in isolated familial DRS pedigree ENG_ET (c.467G>T, p.(Arg156Leu); Supplementary Table 7). The variant is absent from population databases and has damaging predictions, and *OLIG2* is moderately missense-constrained within humans. *Olig2* mouse models have fewer motor neurons and abnormalities of neuronal migration and the hindbrain, while fish models have abnormalities of neuronal migration, CN6, and CN7. Additionally, worm models have abnormal neuronal cell fate specification and axon outgrowth (Supplementary Table 8).

Finally, *SEMA3F*, encoding a semaphorin involved in axon guidance, is mutated in syndromic sporadic CFEOM pedigree ENG_CMK (c.1889C>A, p.(Ser630Ter); Supplementary Table 7). The variant is absent from population databases and has damaging predictions, and *SEMA3F* is LOF-constrained within humans (LOEUF: 0.2190). *Sema3f^-/-^* mice have abnormalities of CN3 and CN4, neuronal migration, and axon guidance and fasciculation. Moreover, worm and fly models have abnormalities of axon guidance and of neuroanatomy and neurophysiology, respectively (Supplementary Table 8). In humans, heterozygous *SEMA3F* missense or LOF variants can result in hypogonadotropic hypogonadism,^37^ which has not been documented in ENG_CMK.

### DenovolyzeR analysis of *de novo* SNVs/indels

After filtering, we identified 297 DNVs (173 missense, 65 synonymous, 30 frameshift, 16 splice site,13 nonsense) among 200 probands. 232 genes had missense, frameshift, splice site, or nonsense variants; 9 genes were mutated twice, but 6 of these were mutated twice in single individuals. Twenty-three genes were not represented in denovolyzeR and were excluded from analysis.

Three genes each had missense DNVs in 2 probands (*TUBA1A, P2RX3*, and *SLC22A6*), but this enrichment did not meet statistical significance after Bonferroni correction for multiple gene testing (p=1.88e-5,1.82e-5, 2.75e-5 for the three genes, respectively; significance threshold at α=0.05 is 1.3e-6). The *TUBA1A* DNV in one of two probands is one of the variants we have reported as causal for the proband’s syndromic CFEOM (pedigree 38; Individual 1^38^), while the second proband had syndromic DRS and has not been reported to date (pedigree 170; c.1136G>A, p.(Ser379Asn), transcript NM_006009.4). While DRS has not been associated with *TUBA1A*, other aberrant innervation patterns have been, and our proband’s syndromic findings are consistent with phenotypes reported in other individuals with *TUBA1A* variants. By contrast, the oCCDD in one of the probands with an *SLC22A6* DNV was solved by another genetic etiology and reported previously (pedigree 144 in our cohort; reported as pedigree 22^39^), and the two individuals with *P2RX3* DNVs have very disparate phenotypes (syndromic CN4-palsy and isolated DRS, respectively), making these less likely to be pathogenic.

### Categorization of genes with ACMG/AMP/ClinGen-classifed SNVs/indels or SVs

The genes which harbored ACMG/AMP/ClinGen-classified P/LP or VUS SNVs/indels or SVs fell into five categories. Among the 14 probands with 14 variants in 7 [oCCDD+,Syndrome+/-] genes *KIF21A, TUBB3, PHOX2A, MAFB, CHN1, SALL4,* and *ROBO3*, five were not prescreened, five were prescreened but had VUS and were sequenced to exclude alternative causes, one was an SV that could not be detected with Sanger sequencing, one had one convincing *ROBO3* allele on pre-screening but no convincing second allele in this gene associated with a recessive condition, and two were missed on prescreening. Among these genes, *KIF21A, TUBB3*, and *ROBO3* had variants in both the P/LP and VUS categories.

The 40 [oCCDD(+),Syndrome+] genes that harbored 61 variants among 56 probands were *PIEZO2, KIAA0586* (compound heterozygous variants in a single proband)*, KIFBP* (compound heterozygous variants in a single proband)*, FGD1, PHOX2B, TRPV4, KMT2D, PTPN11* (mutated in 2 probands)*, ACTB, MED13* (mutated in 2 probands)*, EBF3* (mutated in 3 probands)*, ZC4H2* (mutated in 2 probands)*, BBS1* (compound heterozygous variants in a single proband)*, DMD, HDAC8, GCH1, DYRK1A, ZNF462* (mutated in 4 probands), *TGFBR2* (mutated in 2 probands)*, FBN1, POGZ, ARMC9* (mutated in 2 probands), *ECEL1, COL25A1, KIF21B, TUBB6* (mutated in 2 probands)*, CHD7, OPA1, TOGARAM1* (compound heterozygous variants in a single proband)*, WDR5, MCM3AP* (compound heterozygous variants in a single proband)*, CDC42BPB, TUBB4A, OTUD6B* (compound heterozygous variants in a single proband)*, FLNA, MPZ* (mutated in 2 probands)*, ARX, WDR37, HNRNPK, MYH10* (mutated in 5 probands). Among these genes, *ARMC9*, *PTPN11, ZNF462,* and *TGFBR2* had variants in both the P/LP and VUS categories.

The 13 [oCCDD-,Syndrome+] genes that harbored 18 variants among 17 probands were *ARMC4, SCN1A, CDK13, FOXG1, TGFB2, GJB2, CEP83* (compound heterozygous variants in a single proband)*, TUBA1A* (mutated in 2 probands), *HRAS, COL7A1, SLC12A5, GNAS* (mutated in 4 probands), *MACF1.* Among these genes, *CEP83* and *TUBA1A* had variants in both the P/LP and VUS categories.

The 16 [oCCDD-,Syndrome-] genes that harbored 24 VUS among 24 probands were *TUBA8, TUBA4A* (mutated in 3 probands)*, SEMA3F, OLIG2, FRMD4B, TUBA3E, TUBA1B, TUBB, CTNNA1, KLB, FGF21, FER* (mutated in 2 probands)*, ACTR1B* (mutated in 2 probands)*, KIF5C* (mutated in 3 probands)*, NES, CUX1* (mutated in 3 probands).

The 4 [Misdiagnoses] genes that harbored 5 variants among 5 probands were *FOXL2* (mutated in two probands)*, RYR1, TWIST1*, *CHRNE*. Among these genes, *FOXL2* had a variant in both the P/LP and VUS categories.

### Characteristics of oCCDD probands with ACMG/AMP/ClinGen-P/LP variants

As summarized in Supplementary Table 7, column K, the 13 pedigrees for which ACMG/AMP/ClinGen-P/LP SNV/indels or SVs fully explained the phenotypes are 198, ENG_AWA, ENG_ABW, 81, ENG_JP, 269, 270, 38, ENG_BS, 48, ENG_AKG, ENG_0640, and 193. The 13 pedigrees for which ACMG/AMP/ClinGen-P/LP variant(s) explained the syndromic but not the oCCDD phenotype were 239, ENG_1894, 242, 93, 128, ENG_2270, 42, 227, 238, ENG_AHO, ENG_ADU, ENG_ACR, 257. The 3 pedigrees in which the P/LP variants explained the oCCDD but not the syndromic phenotype were 27, 13, ENG_CMO. The 14 pedigrees in which both the oCCDD and non-CCDD phenotypes were expanded are ENG_BAG, ENG_AGZ, 4, ENG_CKA, ENG_CHA, 61, ENG_FI, 144, 71, 233, 131, 26, 62, ENG_AZW. In two cases, a P/LP allele was compound heterozygous with a VUS in a gene that fully or partially explained the phenotype (pedigrees 193 and ENG_AZW, respectively).

Some VUS had higher levels of supportive evidence and compatibility with prior reported oCCDD and/or syndromic phenotypes consistent with the phenotypes of the probands who harbored them, suggesting a higher likelihood of their being substantiated over time. These included 14 variants in 13 genes among 14 pedigrees: *SALL4* (ENG_DQ), *CHN1* (ENG_1580, ENG_BBG), *MAFB* (232), *PHOX2A* (160), *TUBA1A* (170), *ECEL1* (223), *COL25A1* (56),^40^ *MACF1* (98),^41^ *ARMC9* (ENG_COX), *KIF21B* (ENG_FR), *TUBB6* (ENG_CML), *TUBA4A* (ENG_IM), and *CHRNE* (ENG_2044) (Table 1, Fig. 3H, Supplementary Table 7).

Among oCCDD subphenotypes, ACMG/AMP/ClinGen-P/LP variants were obtained in the following numbers of probands: horizontal gaze palsy=3/6 (50.0%), CCDD-NOS=4/9 (44.4%), CN6-palsy=1/5 (20.0%), CFEOM=14/80 (17.5%), Brown syndrome=2/13 (15.4%), congenital ptosis=7/75 (9.3%), DRS=10/198 (5.1%), CN4-palsy=1/28 (3.6%), and MGJWS=1/53 (1.9%).

Additional breakdowns are provided (Fig. 3I, Supplementary Figure 10).

## SUPPLEMENTARY WEB RESOURCES

http://broadinstitute.github.io/picard/

https://gatk.broadinstitute.org/hc/en-us

https://seqr.broadinstitute.org/

https://github.com/broadinstitute/tgg_methods/

Online Mendelian Inheritance in Man, OMIM®. McKusick-Nathans Institute of Genetic Medicine, Johns Hopkins University (Baltimore, MD). World Wide Web URL: https://omim.org/. Accessed May 2023.

Stenson et al (2003), The Human Gene Mutation Database (HGMD®): 2003 Update. Hum Mutat (2003) 21:577-581. HGMD Professional version 2021.4.

http://monarchinitiative.org

https://github.com/broadinstitute/gatk-sv

## SUPPLEMENTARY FIGURES

**Supplementary Figure 1.**
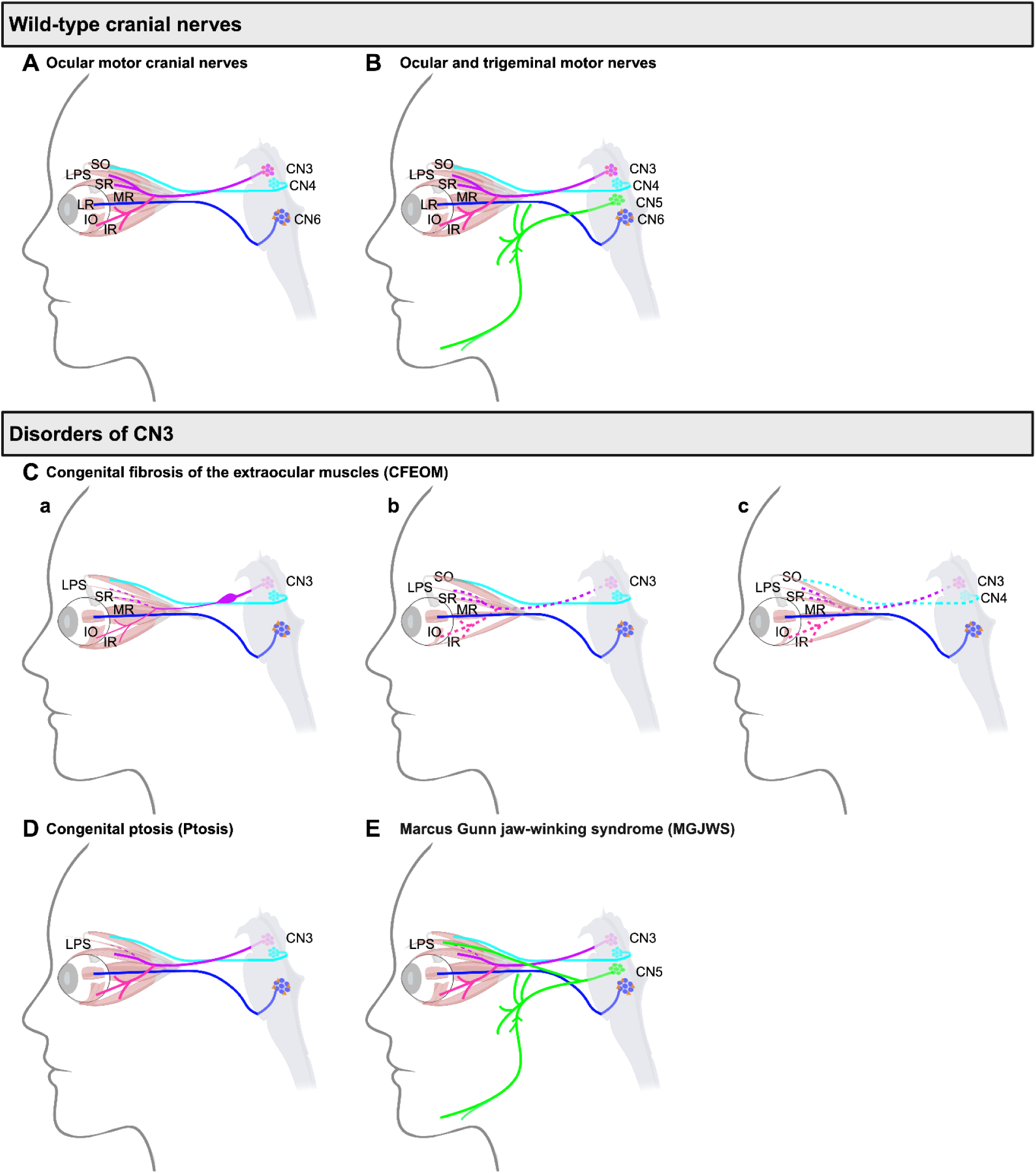
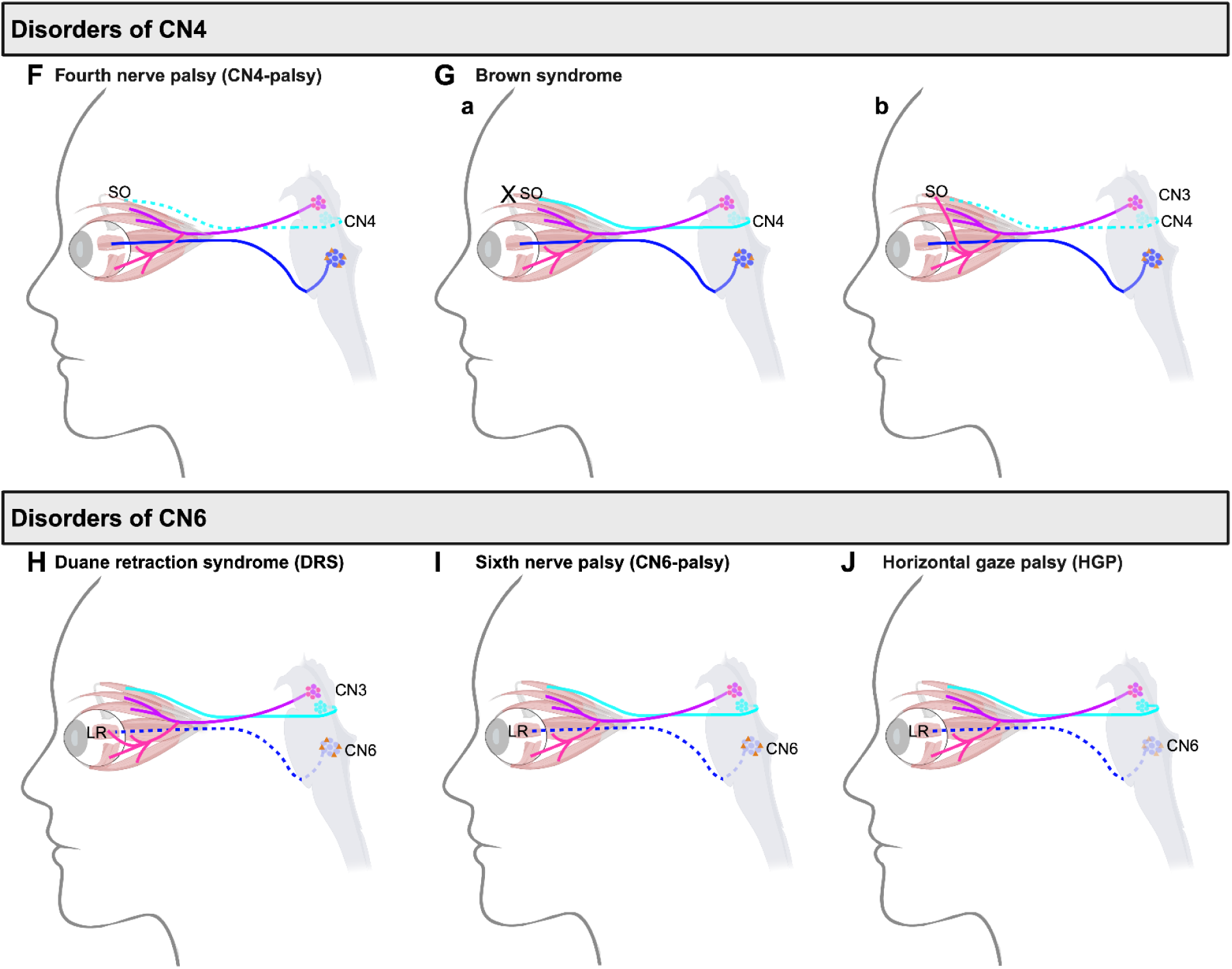
Human cranial nerve schematics. **A:** Wild-type human ocular motor cranial nerves. Three ocular motor cranial motor nuclei (clusters of cells in the brainstem; CN3, CN4, and CN6) give rise to cranial nerves (CN) that innervate 7 target muscles that control eye and/or eyelid movement (IO, IR, LPS, LR, MR, SO, SR). Errors in the identity or migration of these motor neurons or in their axonal growth or guidance can result in oCCDDs. CN3 originates from the midbrain, exits ventrally, and gives rise to a nerve with two main branches: a superior division (purple), which innervates the SR and LPS muscles, and an inferior division (magenta), which innervates the IO, IR, and MR muscles. The CN4 nucleus (cyan) in the midbrain gives rise to the CN4 nerve, which exits the midbrain dorsally, crosses the midline, and passes around the brainstem to innervate the contralateral SO muscle. The CN6 nucleus in the pons contains motor neurons (dark blue) whose axons exit ventrally and innervate the LR muscle, and interneurons (orange) that cross the midline and ascend to contact the medial rectus motor neurons in CN3. **B:** Wild-type human ocular motor cranial nerves are shown as in A, with the addition of the wild-type trigeminal motor nucleus and nerve (CN5), which innervates the muscles of mastication. **C-E:** CN3-related disorders include CFEOM, congenital ptosis, and MGJWS. **C:** CFEOM results from malformation of the superior division of CN3 with corresponding hypoplasia of the SR and LPS muscles (a-c) and can be characterized in mouse models by initial dilation followed by thinning of the nerve (a). CN3 superior division defects may be accompanied by malformation of the inferior division of CN3 (b) with IO, IR, and MR muscle hypoplasia, and, in some cases, malformation of CN4 and the SO muscle (c), or CN6 and the LR muscle (not shown). **D:** Congenital ptosis can be caused by maldevelopment of the superior branch of CN3 to the LPS with corresponding LPS hypoplasia. **E:** In congenital ptosis accompanied by MGJWS, the superior branch of CN3 to the LPS is deficient as in congenital ptosis, but the LPS is aberrantly innervated by CN5. The mechanisms and precise branches of CN5 involved in this phenotype are poorly understood. **F-G:** CN4-related disorders include CN4-palsy and Brown syndrome. **F:** In CN4-palsy, CN4 and its SO target muscle are hypoplastic. **G:** Brown syndrome can result from limited motility of the SO muscle or its tendon sheath (a). Some cases of Brown syndrome are alternatively hypothesized to be oCCDDs resulting from CN4-palsy with aberrant innervation, potentially from the inferior division of CN3 (b), but such mechanisms are not proven. **H-J:** CN6-related disorders include DRS, CN6-palsy, and HGP. **H:** DRS is characterized by CN6 motor neuron maldevelopment or axon stalling, with secondary aberrant innervation of the LR muscle by the inferior division of CN3. In DRS, the CN6 motor nucleus interneurons (orange) are spared. **I:** Congenital CN6-palsy arises secondary to CN6 maldevelopment or degeneration, with sparing of the interneurons. **J:** HGP can be caused by abnormalities of CN6, including its interneurons, or the brain regions that project to CN6, such as the medial longitudinal fasciculus or paramedian pontine reticular formation (not shown). Abbreviations: CFEOM=congenital fibrosis of the extraocular muscles, CN=cranial nerve, CN3=cranial nerve 3 (oculomotor), CN4=cranial nerve 4 (trochlear), CN4-palsy=fourth nerve palsy, CN5=cranial nerve 5 (trigeminal motor), CN6=cranial nerve 6 (abducens), CN6-palsy=sixth nerve palsy, DRS=Duane retraction syndrome, HGP=horizontal gaze palsy, IO=inferior oblique muscle, IR=inferior rectus muscle, LPS=levator palpebrae superioris muscle, LR=lateral rectus muscle, MGJWS=Marcus Gunn jaw-winking syndrome, MR=medial rectus muscle, SO=superior oblique muscle, SR=superior rectus muscle. Key: purple=CN3 superior division, pink=CN3 inferior division, cyan=CN4, green=CN5, dark blue=CN6, orange=CN6 interneurons, “X” or increased transparency=structure/function compromised, dashed nerve=nerve missing or hypoplastic.

**Supplementary Figure 2.**
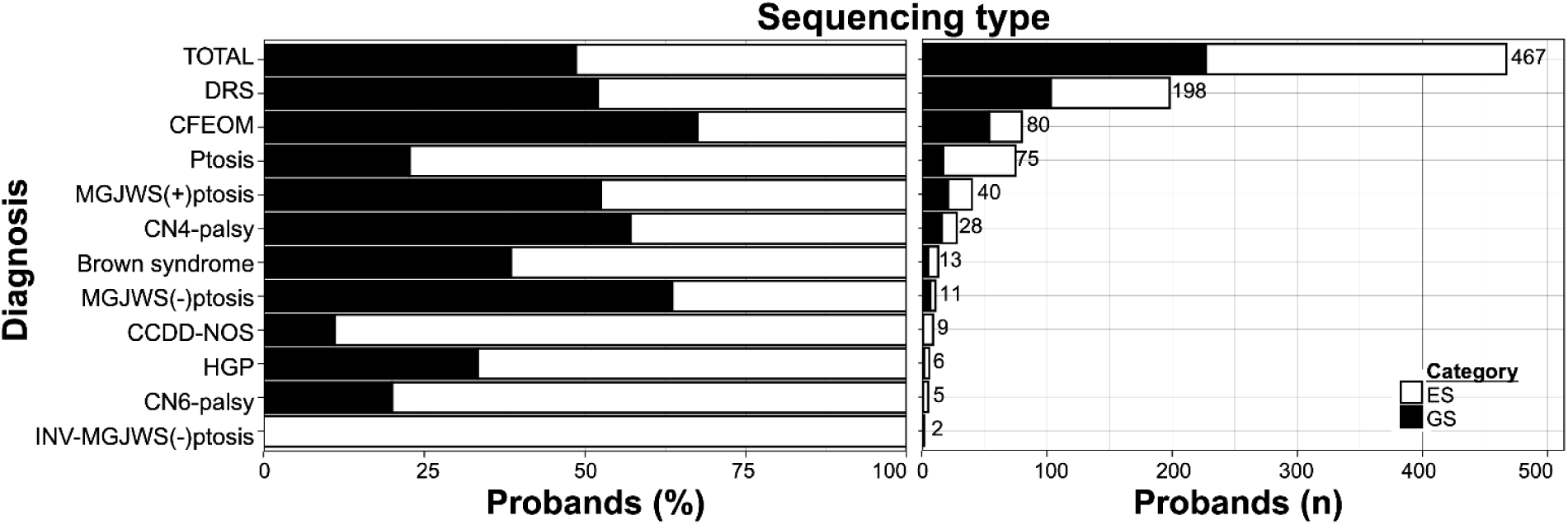
Sequencing type among 467 probands. Abbreviations: CCDD=congenital cranial dysinnervation disorder, CCDD-NOS=CCDD not otherwise specified, CFEOM=congenital fibrosis of the extraocular muscles, CN4-palsy=fourth nerve palsy, CN6-palsy=sixth nerve palsy, DRS=Duane retraction syndrome, ES=exome sequencing, GS=genome sequencing, HGP=horizontal gaze palsy, INV-MGJWS(-)ptosis=inverse Marcus Gunn jaw-winking synkinesis without congenital ptosis, MGJWS(+)ptosis=Marcus Gunn jaw-winking synkinesis with congenital ptosis, MGJWS(-)ptosis=Marcus Gunn jaw-winking synkinesis without congenital ptosis, oCCDD=ocular congenital cranial dysinnervation disorder, Ptosis=congenital ptosis.

**Supplementary Figure 3.**
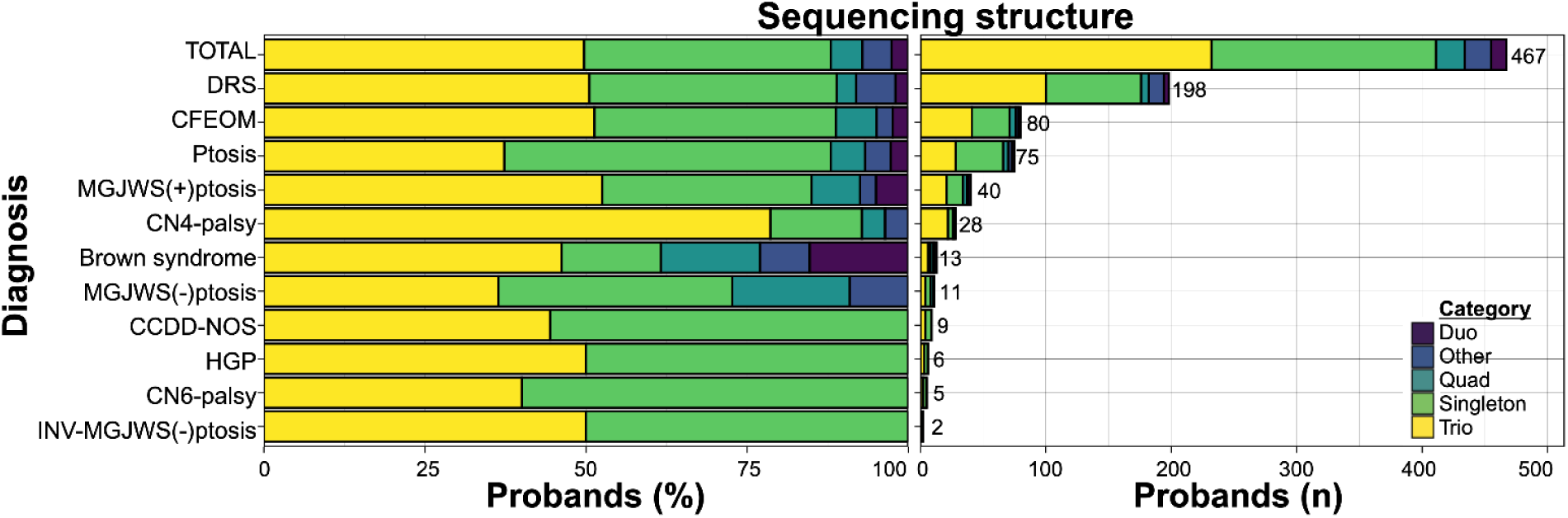
Sequencing structure among 467 probands. Abbreviations: CCDD=congenital cranial dysinnervation disorder, CCDD-NOS=CCDD not otherwise specified, CFEOM=congenital fibrosis of the extraocular muscles, CN4-palsy=fourth nerve palsy, CN6-palsy=sixth nerve palsy, DRS=Duane retraction syndrome, Duo=2 members of pedigree sequenced, HGP=horizontal gaze palsy, INV-MGJWS(-)ptosis=inverse Marcus Gunn jaw-winking synkinesis without congenital ptosis, MGJWS(+)ptosis=Marcus Gunn jaw-winking synkinesis with congenital ptosis, MGJWS(-)ptosis=Marcus Gunn jaw-winking synkinesis without congenital ptosis, oCCDD=ocular congenital cranial dysinnervation disorder, Other= more than 4 members of pedigree sequenced, Ptosis=congenital ptosis, Quad=4 members of pedigree sequenced, Singleton=1 member of pedigree sequenced, Trio=3 members of pedigree sequenced.

**Supplementary Figure 4.**
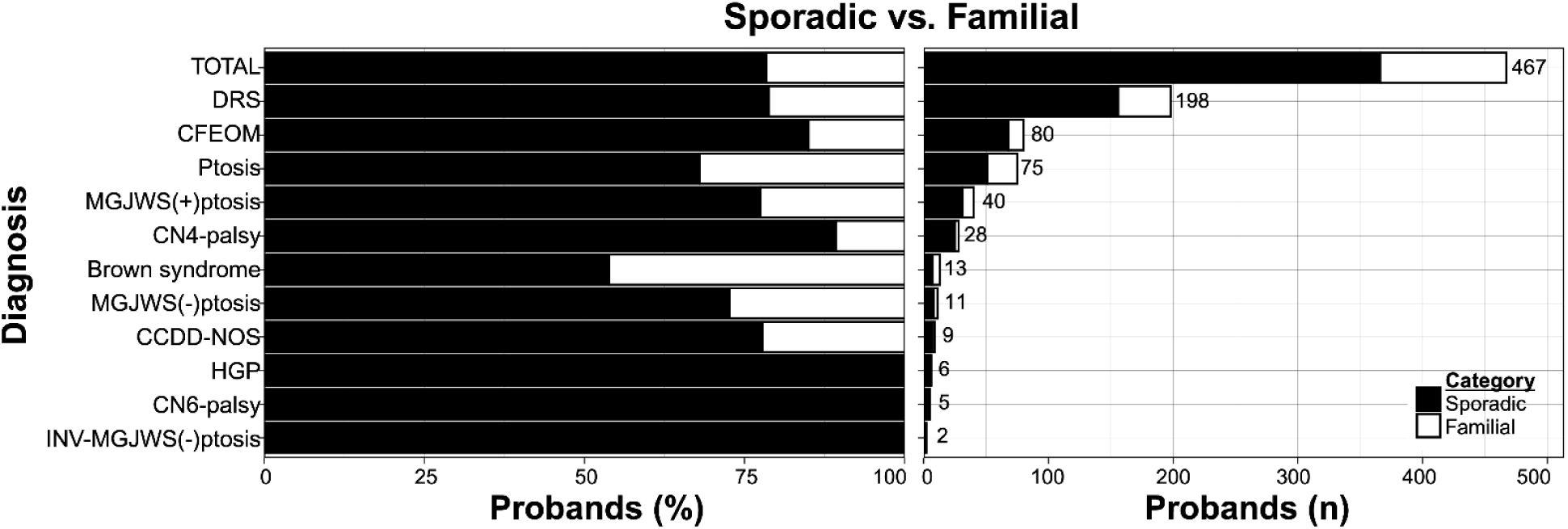
Sporadic versus familial oCCDDs among 467 probands. Abbreviations: CCDD=congenital cranial dysinnervation disorder, CCDD-NOS=CCDD not otherwise specified, CFEOM=congenital fibrosis of the extraocular muscles, CN4-palsy=fourth nerve palsy, CN6-palsy=sixth nerve palsy, DRS=Duane retraction syndrome, HGP=horizontal gaze palsy, INV-MGJWS(-)ptosis=inverse Marcus Gunn jaw-winking synkinesis without congenital ptosis, MGJWS(+)ptosis=Marcus Gunn jaw-winking synkinesis with congenital ptosis, MGJWS(-)ptosis=Marcus Gunn jaw-winking synkinesis without congenital ptosis, oCCDD=ocular congenital cranial dysinnervation disorder, Ptosis=congenital ptosis.

**Supplementary Figure 5.**
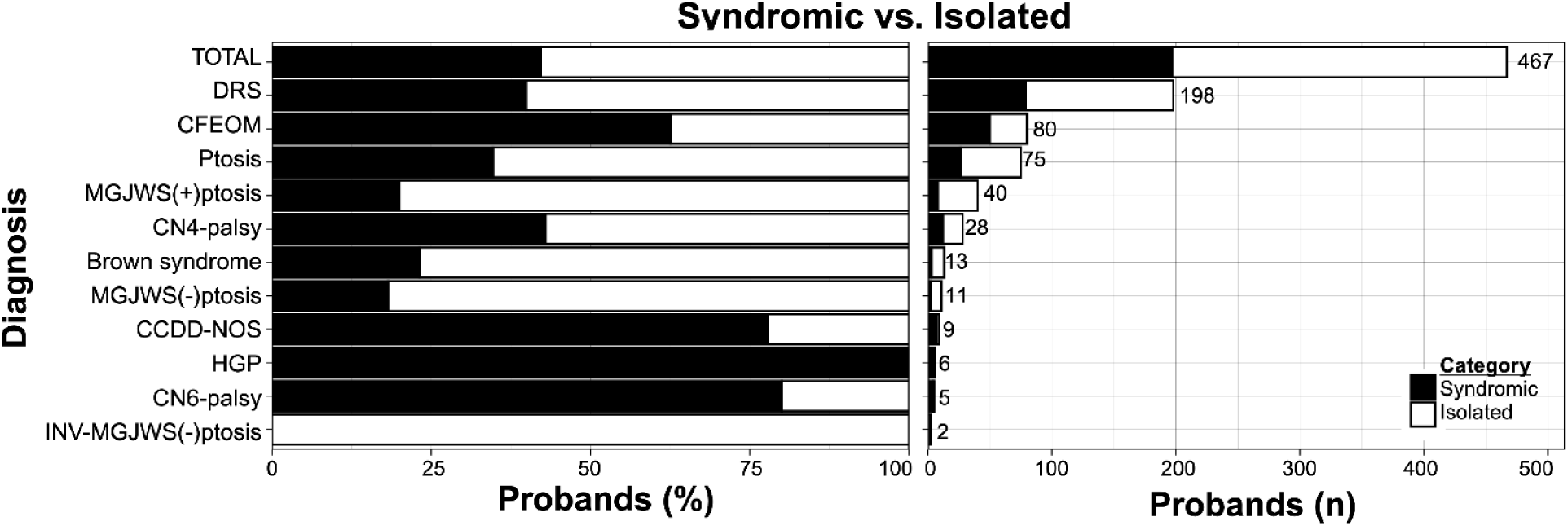
Syndromic versus isolated oCCDD breakdown among 467 probands. Abbreviations: CCDD=congenital cranial dysinnervation disorder, CCDD-NOS=CCDD not otherwise specified, CFEOM=congenital fibrosis of the extraocular muscles, CN4-palsy=fourth nerve palsy, CN6-palsy=sixth nerve palsy, DRS=Duane retraction syndrome, HGP=horizontal gaze palsy, INV-MGJWS(-)ptosis=inverse Marcus Gunn jaw-winking synkinesis without congenital ptosis, MGJWS(+)ptosis=Marcus Gunn jaw-winking synkinesis with congenital ptosis, MGJWS(-)ptosis=Marcus Gunn jaw-winking synkinesis without congenital ptosis, oCCDD=ocular congenital cranial dysinnervation disorder, Ptosis=congenital ptosis.

**Supplementary Figure 6.**
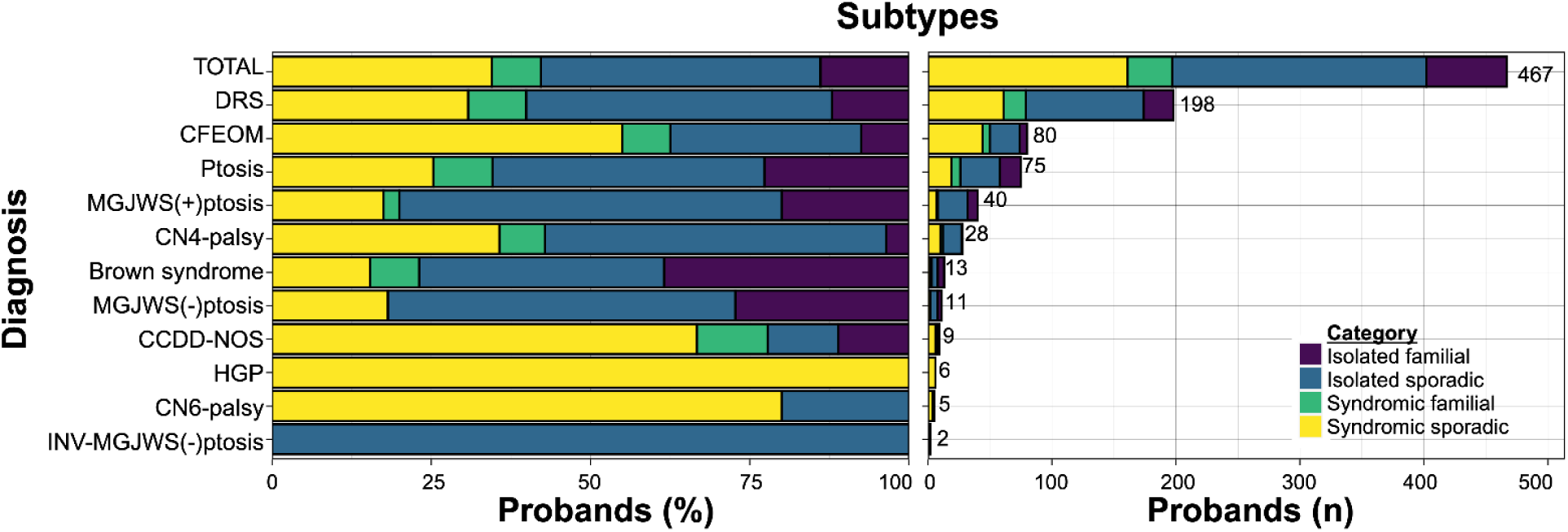
Combined sporadic/familial and isolated/syndromic oCCDD breakdown among 467 probands. Abbreviations: CCDD=congenital cranial dysinnervation disorder, CCDD-NOS=CCDD not otherwise specified, CFEOM=congenital fibrosis of the extraocular muscles, CN4-palsy=fourth nerve palsy, CN6-palsy=sixth nerve palsy, DRS=Duane retraction syndrome, HGP=horizontal gaze palsy, INV-MGJWS(-)ptosis=inverse Marcus Gunn jaw-winking synkinesis without congenital ptosis, MGJWS(+)ptosis=Marcus Gunn jaw-winking synkinesis with congenital ptosis, MGJWS(-)ptosis=Marcus Gunn jaw-winking synkinesis without congenital ptosis, oCCDD=ocular congenital cranial dysinnervation disorder, Ptosis=congenital ptosis.

**Supplementary Figure 7.**
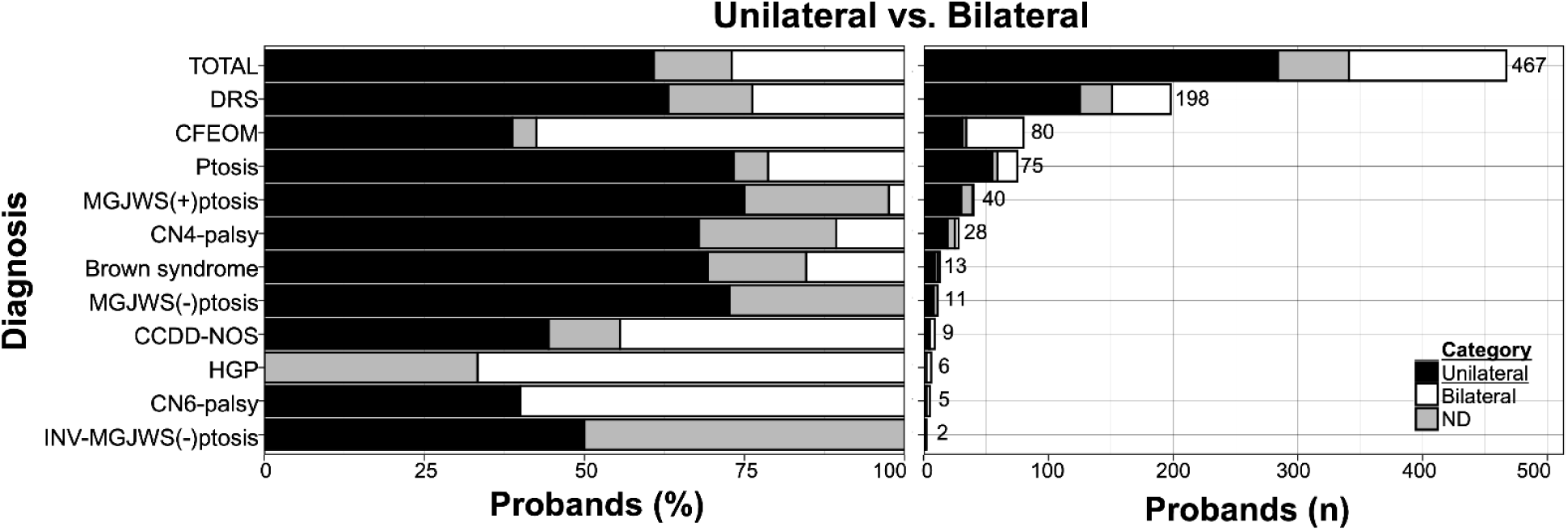
Laterality of oCCDDs among 467 probands. Abbreviations: CCDD=congenital cranial dysinnervation disorder, CCDD-NOS=CCDD not otherwise specified, CFEOM=congenital fibrosis of the extraocular muscles, CN4-palsy=fourth nerve palsy, CN6-palsy=sixth nerve palsy, DRS=Duane retraction syndrome, HGP=horizontal gaze palsy, INV-MGJWS(-)ptosis=inverse Marcus Gunn jaw-winking synkinesis without congenital ptosis, MGJWS(+)ptosis=Marcus Gunn jaw-winking synkinesis with congenital ptosis, MGJWS(-)ptosis=Marcus Gunn jaw-winking synkinesis without congenital ptosis, ND=not described, oCCDD=ocular congenital cranial dysinnervation disorder, Ptosis=congenital ptosis.

**Supplementary Figure 8.**
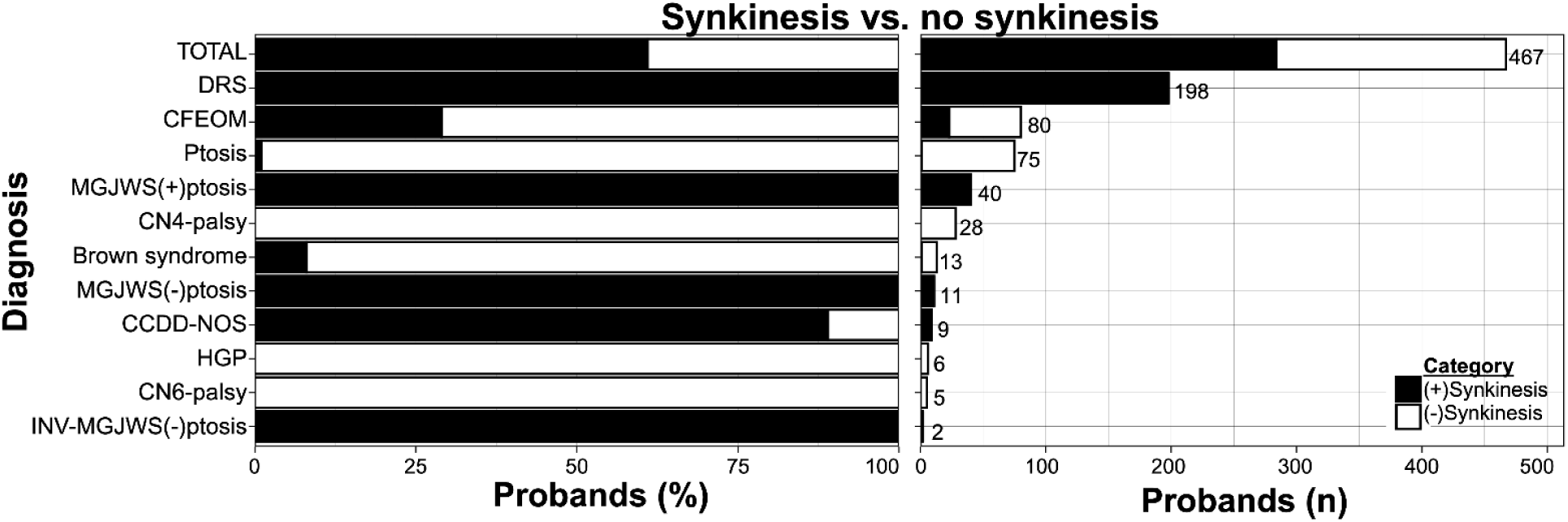
Documented synkinesis among 467 probands. Abbreviations: CCDD=congenital cranial dysinnervation disorder, CCDD-NOS=CCDD not otherwise specified, CFEOM=congenital fibrosis of the extraocular muscles, CN4-palsy=fourth nerve palsy, CN6-palsy=sixth nerve palsy, DRS=Duane retraction syndrome, HGP=horizontal gaze palsy, INV-MGJWS(-)ptosis=inverse Marcus Gunn jaw-winking synkinesis without congenital ptosis, MGJWS(+)ptosis=Marcus Gunn jaw-winking synkinesis with congenital ptosis, MGJWS(-)ptosis=Marcus Gunn jaw-winking synkinesis without congenital ptosis, ND=not described, oCCDD=ocular congenital cranial dysinnervation disorder, Ptosis=congenital ptosis.

**Supplementary Figure 9.**
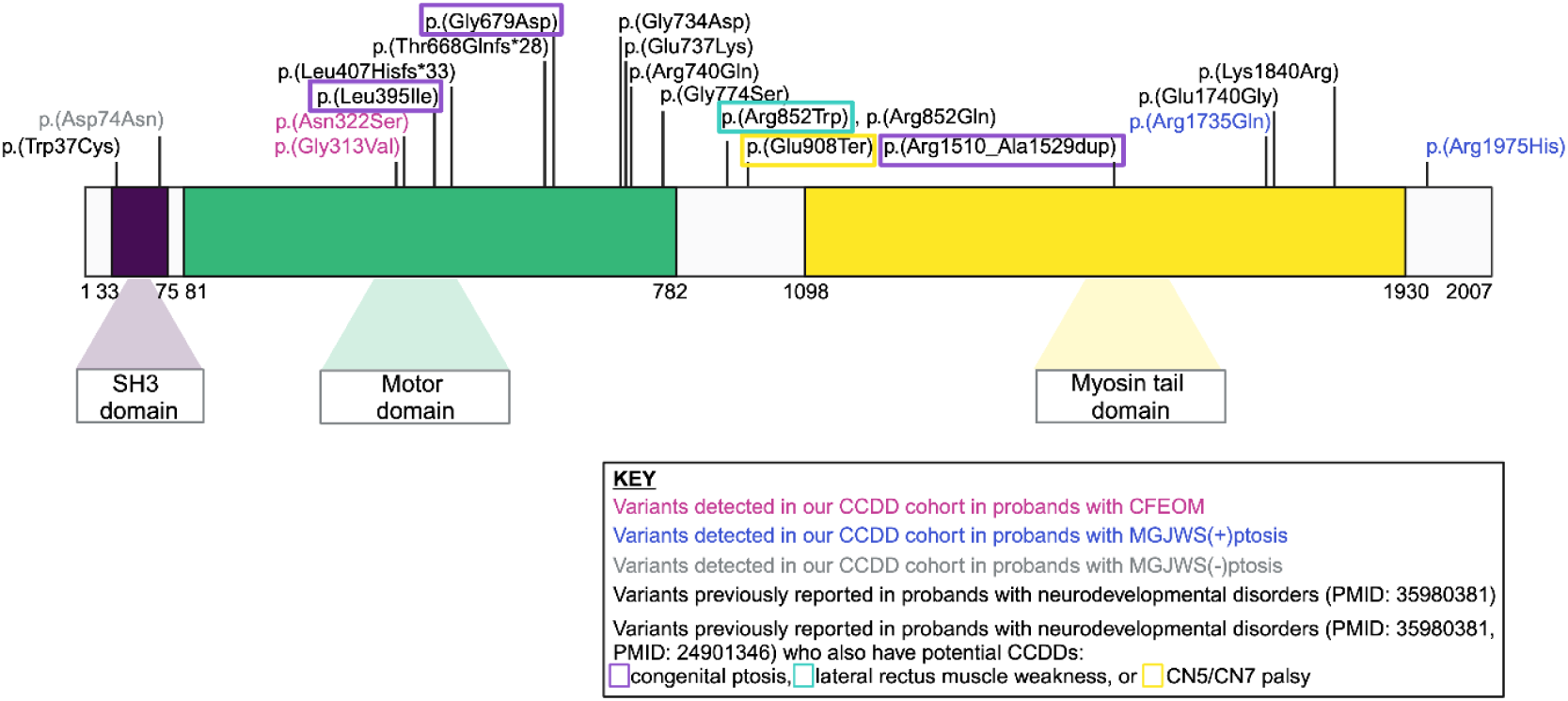
Protein mapping of *MYH10* variants identified in our cohort and reported in the literature. Abbreviations: CCDD=congenital cranial dysinnervation disorder, CFEOM=congenital fibrosis of the extraocular muscles, MGJWS(+)ptosis=Marcus Gunn jaw-winking synkinesis with congenital ptosis, MGJWS(-)ptosis=Marcus Gunn jaw-winking synkinesis without congenital ptosis, Ptosis=congenital ptosis. Variants mapped with ENST00000360416.8 (NM_001256012.3).

**Supplementary Figure 10.**
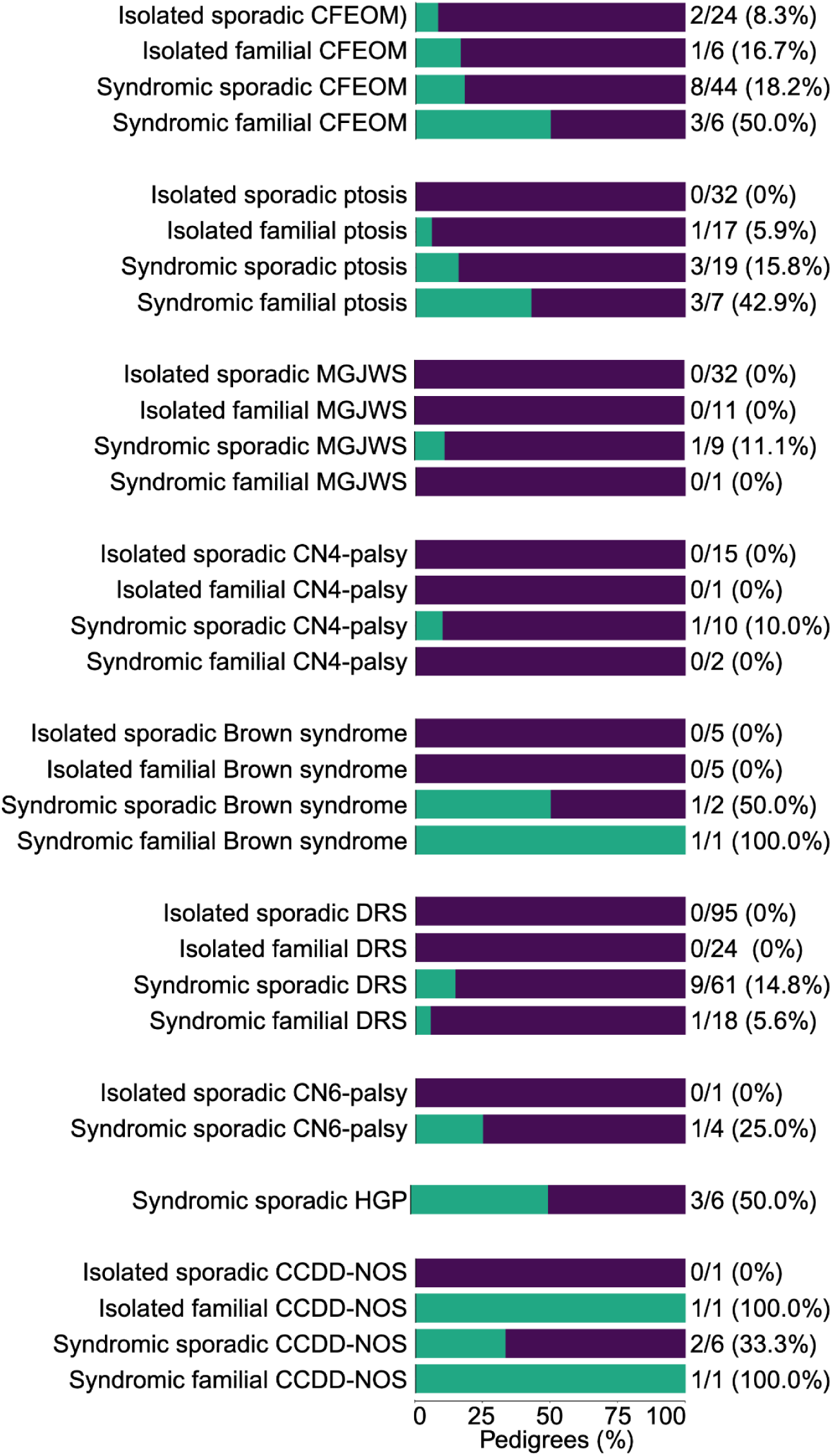
Characteristics among pedigrees with ACMG/AMP/ClinGen-P/LP variants. Key: green-counts of probands with ACMG/AMP/ClinGen-P/LP variants, purple-counts of probands without ACMG/AMP/ClinGen-P/LP variants. Abbreviations: ACMG=American College of Medical Genetics and Genomics, AMP=Association for Molecular Pathology, CCDD=congenital cranial dysinnervation disorder, CCDD-NOS=CCDD not otherwise specified, CFEOM=congenital fibrosis of the extraocular muscles, ClinGen=Clinical Genome Resource, CN4-palsy=fourth nerve palsy, CN6-palsy=sixth nerve palsy, DRS=Duane retraction syndrome, HGP=horizontal gaze palsy, LP=likely pathogenic ACMG/AMP/ClinGen classification, MGJWS=Marcus Gunn jaw-winking synkinesis, oCCDD=ocular congenital cranial dysinnervation disorder, P=pathogenic ACMG/AMP/ClinGen classification, Ptosis=congenital ptosis.

## SUPPLEMENTARY TABLES

**Supplementary Table 2.**
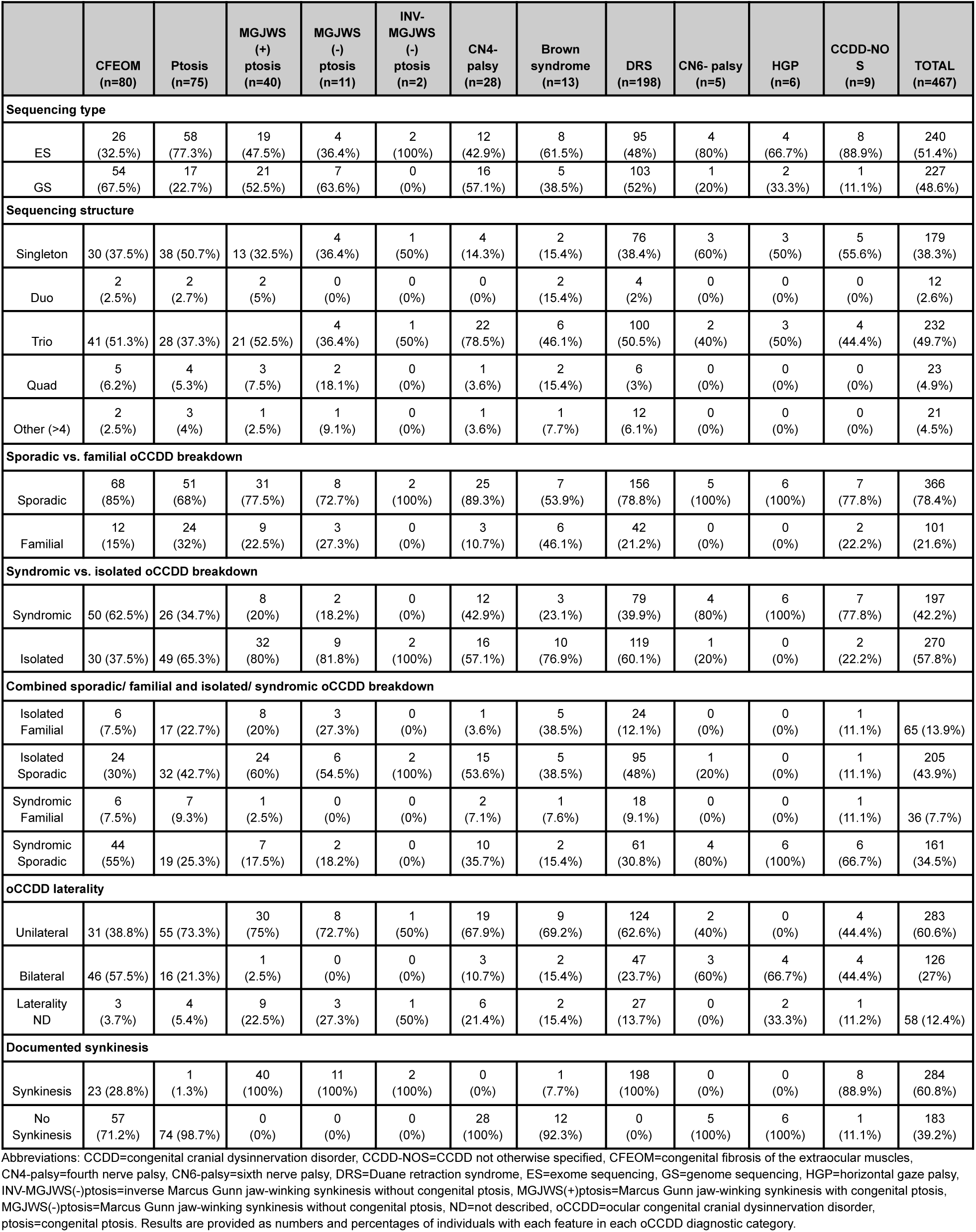
Sequencing, demographics, and oCCDD information among 467 probands.

**Supplementary Table 3.**
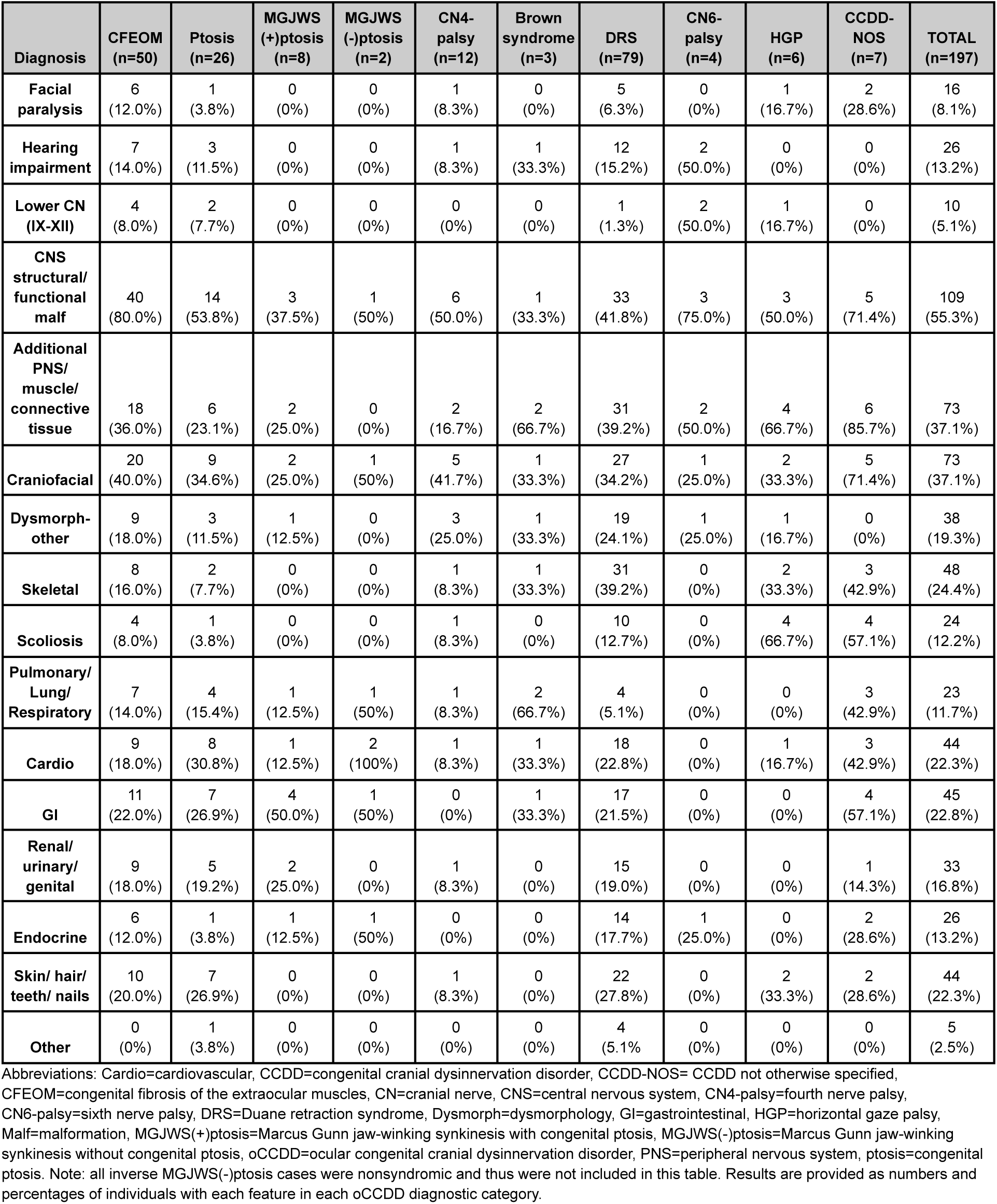
Syndromic features among the 197 probands with syndromic oCCDDs.

**Supplementary Table 4.** CODA analysis of co-occurring syndromic phenotypes.

| oCCDD<br>Diagnosis | Defect A | Defect B | Defect C | Defect D | Defect E | Number of<br>probands with<br>phenotype<br>combination | OEun | OEadj |
| --- | --- | --- | --- | --- | --- | --- | --- | --- |
| DRS | 4 | 5 | 6 | 7 | 8 | 4 | 9.58 | 5.30 |
| DRS | 4 | 5 | 6 | 7 | 9 | 4 | 29.69 | 11.39 |
| DRS | 4 | 5 | 6 | 7 | 15 | 3 | 10.12 | 5.86 |
| DRS | 4 | 5 | 6 | 8 | 9 | 3 | 13.65 | 6.11 |
| DRS | 4 | 5 | 6 | 8 | 10 | 3 | 34.12 | 15.08 |
| DRS | 4 | 5 | 6 | 9 | 12 | 3 | 24.89 | 8.75 |
| DRS | 4 | 5 | 8 | 9 | 12 | 3 | 21.67 | 12.02 |
| DRS | 5 | 6 | 7 | 8 | 15 | 3 | 10.77 | 6.76 |
| DRS | 5 | 6 | 7 | 12 | 15 | 3 | 19.65 | 10.72 |
| DRS | 6 | 7 | 8 | 12 | 15 | 3 | 19.65 | 8.94 |
| DRS | 6 | 7 | 9 | 12 | 15 | 3 | 60.90 | 17.42 |
| DRS | 4 | 6 | 7 | 8 | 14 | 3 | 15.90 | 8.83 |
| DRS | 1 | 4 | 5 | 15 |  | 3 | 13.14 | 6.08 |
| DRS | 4 | 5 | 6 | 12 |  | 4 | 4.20 | 3.13 |
| DRS | 4 | 5 | 6 | 15 |  | 5 | 4.06 | 3.02 |
| DRS | 4 | 5 | 8 | 15 |  | 3 | 2.12 | 1.85 |
| DRS | 4 | 5 | 9 | 12 |  | 4 | 11.34 | 7.77 |
| DRS | 4 | 5 | 10 | 12 |  | 3 | 21.26 | 12.57 |
| DRS | 4 | 5 | 12 | 15 |  | 3 | 3.87 | 2.47 |
| DRS | 4 | 6 | 7 | 8 |  | 6 | 5.64 | 5.15 |
| DRS | 4 | 6 | 7 | 12 |  | 3 | 5.14 | 3.53 |
| DRS | 4 | 6 | 8 | 12 |  | 3 | 3.15 | 2.68 |
| DRS | 5 | 6 | 8 | 12 |  | 3 | 3.35 | 2.92 |
| DRS | 5 | 7 | 8 | 15 |  | 4 | 4.91 | 5.14 |
| DRS | 6 | 7 | 8 | 9 |  | 3 | 9.30 | 4.05 |
| DRS | 6 | 7 | 8 | 12 |  | 4 | 7.29 | 4.76 |
| DRS | 6 | 8 | 9 | 12 |  | 3 | 10.40 | 6.01 |
| DRS | 7 | 8 | 9 | 12 |  | 3 | 14.77 | 6.42 |
| DRS | 4 | 5 | 6 | 13 |  | 3 | 3.57 | 2.46 |
| DRS | 4 | 5 | 9 | 13 |  | 3 | 9.64 | 6.04 |
| DRS | 4 | 5 | 13 | 15 |  | 3 | 4.38 | 2.96 |
| DRS | 5 | 6 | 12 | 13 |  | 3 | 6.93 | 6.59 |
| DRS | 5 | 6 | 8 | 13 |  | 4 | 5.07 | 4.83 |
| DRS | 6 | 7 | 8 | 11 |  | 3 | 5.17 | 3.00 |
| DRS | 6 | 8 | 11 | 13 |  | 4 | 8.73 | 6.72 |
| DRS | 2 | 4 | 5 | 6 |  | 4 | 5.95 | 4.45 |
| DRS | 2 | 4 | 6 | 8 |  | 3 | 4.46 | 3.94 |
| DRS | 4 | 6 | 7 | 11 |  | 3 | 4.85 | 3.78 |
| DRS | 4 | 6 | 8 | 11 |  | 3 | 2.98 | 2.30 |
| DRS | 4 | 8 | 11 | 14 |  | 3 | 5.74 | 8.98 |
| DRS | 4 | 5 | 6 | 14 |  | 3 | 3.83 | 2.53 |
| DRS | 2 | 4 | 11 |  |  | 3 | 2.63 | 3.34 |
| DRS | 2 | 4 | 12 |  |  | 3 | 2.78 | 2.92 |
| DRS | 4 | 11 | 12 |  |  | 4 | 2.47 | 2.86 |
| DRS | 1 | 4 | 5 |  |  | 4 | 4.88 | 4.15 |
| DRS | 4 | 12 | 15 |  |  | 4 | 2.02 | 2.13 |
| DRS | 5 | 7 | 9 |  |  | 5 | 5.30 | 4.87 |
| DRS | 5 | 7 | 12 |  |  | 4 | 2.49 | 2.52 |
| DRS | 5 | 8 | 9 |  |  | 4 | 2.60 | 3.06 |
| DRS | 5 | 9 | 15 |  |  | 3 | 2.75 | 2.11 |
| DRS | 7 | 9 | 12 |  |  | 4 | 7.73 | 5.57 |
| DRS | 6 | 7 | 13 |  |  | 3 | 2.43 | 2.18 |
| DRS | 4 | 5 | 11 |  |  | 4 | 1.36 | 1.38 |
| DRS | 5 | 6 | 11 |  |  | 4 | 1.66 | 1.79 |
| DRS | 5 | 13 | 15 |  |  | 4 | 2.44 | 2.53 |
| DRS | 6 | 13 | 15 |  |  | 3 | 2.10 | 2.21 |
| DRS | 11 | 12 | 14 |  |  | 3 | 4.37 | 4.00 |
| DRS | 5 | 8 | 11 |  |  | 3 | 1.08 | 1.42 |
| DRS | 5 | 11 | 12 |  |  | 3 | 1.97 | 2.17 |
| DRS | 5 | 11 | 15 |  |  | 3 | 1.53 | 1.66 |
| DRS | 8 | 11 | 12 |  |  | 3 | 1.97 | 2.17 |
| DRS | 4 | 5 | 14 |  |  | 5 | 2.18 | 2.30 |
| DRS | 4 | 12 | 14 |  |  | 4 | 3.18 | 3.17 |
| DRS | 8 | 12 | 14 |  |  | 3 | 2.54 | 2.96 |
| DRS | 4 | 14 | 15 |  |  | 3 | 1.84 | 2.05 |
| DRS | 2 | 6 | 14 |  |  | 3 | 4.13 | 4.55 |
| DRS | 4 | 15 |  |  |  | 13 | 1.41 | 2.16 |
| DRS | 9 | 15 |  |  |  | 4 | 1.44 | 1.84 |
| DRS | 2 | 13 |  |  |  | 3 | 1.32 | 1.78 |
| DRS | 11 | 14 |  |  |  | 5 | 1.57 | 1.93 |
| DRS | 7 | 11 |  |  |  | 5 | 1.15 | 1.42 |
| DRS | 8 | 11 |  |  |  | 8 | 1.13 | 1.59 |
| DRS | 8 | 13 |  |  |  | 7 | 1.19 | 1.71 |
| DRS | 11 | 13 |  |  |  | 5 | 1.46 | 1.79 |
| DRS | 6 | 14 |  |  |  | 6 | 1.25 | 1.61 |
| DRS | 8 | 14 |  |  |  | 5 | 0.91 | 1.24 |
| CFEOM | 4 | 5 | 6 | 7 | 8 | 4 | 24.11 | 12.22 |
| CFEOM | 4 | 5 | 6 | 7 | 15 | 3 | 14.47 | 12.81 |
| CFEOM | 4 | 5 | 6 | 8 | 15 | 3 | 16.28 | 7.62 |
| CFEOM | 4 | 5 | 7 | 8 | 15 | 3 | 36.17 | 15.63 |
| CFEOM | 4 | 6 | 7 | 8 | 15 | 3 | 32.55 | 12.33 |
| CFEOM | 5 | 6 | 7 | 8 | 15 | 3 | 72.34 | 16.56 |
| CFEOM | 3 | 4 | 5 | 6 | 10 | 3 | 46.50 | 19.63 |
| CFEOM | 3 | 4 | 5 | 6 | 14 | 3 | 54.25 | 16.42 |
| CFEOM | 3 | 4 | 5 | 10 | 14 | 3 | 155.01 | 32.00 |
| CFEOM | 3 | 4 | 6 | 10 | 14 | 3 | 139.51 | 31.73 |
| CFEOM | 3 | 5 | 6 | 10 | 14 | 3 | 310.02 | 33.25 |
| CFEOM | 4 | 5 | 6 | 8 | 14 | 3 | 27.13 | 9.05 |
| CFEOM | 4 | 5 | 6 | 10 | 14 | 3 | 31.00 | 16.50 |
| CFEOM | 3 | 4 | 5 | 6 | 12 | 3 | 29.59 | 17.68 |
| CFEOM | 4 | 5 | 6 | 13 |  | 3 | 2.89 | 3.30 |
| CFEOM | 4 | 6 | 7 | 13 |  | 3 | 5.79 | 4.95 |
| CFEOM | 4 | 6 | 8 | 13 |  | 3 | 6.51 | 4.67 |
| CFEOM | 4 | 6 | 8 | 15 |  | 4 | 7.81 | 7.50 |
| CFEOM | 2 | 4 | 5 | 6 |  | 3 | 3.72 | 3.73 |
| CFEOM | 2 | 4 | 6 | 10 |  | 3 | 9.57 | 6.76 |
| CFEOM | 2 | 4 | 6 | 14 |  | 3 | 11.16 | 7.21 |
| CFEOM | 3 | 4 | 5 | 6 |  | 4 | 8.68 | 9.51 |
| CFEOM | 4 | 5 | 6 | 14 |  | 4 | 5.79 | 6.60 |
| CFEOM | 4 | 5 | 8 | 10 |  | 3 | 9.30 | 6.53 |
| CFEOM | 4 | 5 | 6 | 11 |  | 3 | 2.89 | 3.02 |
| CFEOM | 4 | 5 | 8 | 11 |  | 3 | 7.23 | 5.28 |
| CFEOM | 4 | 5 | 11 | 12 |  | 3 | 5.26 | 6.06 |
| CFEOM | 4 | 6 | 7 | 11 |  | 3 | 5.79 | 5.57 |
| CFEOM | 4 | 6 | 10 | 12 |  | 4 | 8.12 | 8.82 |
| CFEOM | 4 | 6 | 11 | 12 |  | 3 | 4.73 | 4.62 |
| CFEOM | 4 | 6 | 11 | 14 |  | 3 | 8.68 | 7.56 |
| CFEOM | 4 | 6 | 11 | 13 |  | 3 | 5.79 | 5.57 |
| CFEOM | 4 | 6 | 13 | 14 |  | 3 | 8.68 | 6.61 |
| CFEOM | 4 | 6 | 12 | 13 |  | 3 | 4.73 | 4.62 |
| CFEOM | 4 | 6 | 13 |  |  | 6 | 2.08 | 3.49 |
| CFEOM | 4 | 6 | 15 |  |  | 5 | 1.56 | 2.43 |
| CFEOM | 5 | 6 | 13 |  |  | 4 | 3.09 | 3.33 |
| CFEOM | 1 | 4 | 5 |  |  | 3 | 1.74 | 2.32 |
| CFEOM | 1 | 4 | 6 |  |  | 3 | 1.56 | 2.13 |
| CFEOM | 2 | 4 | 6 |  |  | 6 | 2.68 | 4.08 |
| CFEOM | 3 | 4 | 5 |  |  | 4 | 3.47 | 5.09 |
| CFEOM | 3 | 4 | 6 |  |  | 4 | 3.13 | 4.33 |
| CFEOM | 4 | 5 | 10 |  |  | 4 | 1.98 | 2.79 |
| CFEOM | 4 | 6 | 10 |  |  | 5 | 2.23 | 3.20 |
| CFEOM | 4 | 6 | 14 |  |  | 5 | 2.60 | 4.03 |
| CFEOM | 4 | 5 | 11 |  |  | 5 | 1.93 | 2.99 |
| CFEOM | 4 | 5 | 12 |  |  | 5 | 1.58 | 2.26 |
| CFEOM | 4 | 6 | 11 |  |  | 5 | 1.74 | 2.58 |
| CFEOM | 4 | 6 | 12 |  |  | 6 | 1.70 | 2.49 |
| CFEOM | 4 | 7 | 12 |  |  | 3 | 1.89 | 2.27 |
| CFEOM | 4 | 9 | 12 |  |  | 3 | 4.26 | 4.99 |
| CFEOM | 4 | 10 | 11 |  |  | 4 | 3.97 | 5.07 |
| CFEOM | 4 | 12 | 15 |  |  | 4 | 2.27 | 2.88 |
| CFEOM | 2 | 4 | 7 |  |  | 3 | 2.98 | 3.80 |
| CFEOM | 1 | 4 | 12 |  |  | 3 | 2.84 | 3.26 |
| CFEOM | 2 | 4 | 12 |  |  | 3 | 2.44 | 2.59 |
| CFEOM | 1 | 5 |  |  |  | 4 | 1.85 | 2.34 |
| CFEOM | 4 | 9 |  |  |  | 4 | 1.25 | 1.90 |
| Congenital ptosis | 2 | 4 | 5 | 15 |  | 2 | 19.93 | 18.57 |
| Congenital ptosis | 4 | 5 | 10 | 13 |  | 2 | 20.92 | 17.89 |
| Congenital ptosis | 4 | 10 | 11 | 12 |  | 2 | 11.21 | 15.32 |
| Congenital ptosis | 2 | 4 | 6 | 12 |  | 2 | 13.28 | 11.42 |
| Congenital ptosis | 3 | 4 | 5 | 12 |  | 2 | 29.89 | 22.90 |
| Congenital ptosis | 4 | 6 | 11 |  |  | 2 | 1.34 | 1.78 |
| Congenital ptosis | 2 | 4 | 13 |  |  | 2 | 6.44 | 6.29 |
| Congenital ptosis | 4 | 5 | 13 |  |  | 3 | 4.83 | 5.70 |
| Congenital ptosis | 4 | 6 | 12 |  |  | 3 | 2.30 | 3.63 |
| Congenital ptosis | 4 | 11 | 12 |  |  | 3 | 2.59 | 3.78 |
| Congenital ptosis | 4 | 12 | 13 |  |  | 2 | 2.76 | 3.08 |
| Congenital ptosis | 4 | 6 | 15 |  |  | 2 | 1.53 | 2.42 |
| Congenital ptosis | 6 | 7 | 15 |  |  | 2 | 7.15 | 10.18 |
| Congenital ptosis | 4 | 5 | 11 |  |  | 2 | 2.01 | 2.36 |
| Congenital ptosis | 6 | 11 |  |  |  | 3 | 1.08 | 1.67 |
| Congenital ptosis | 2 | 4 |  |  |  | 3 | 1.86 | 2.87 |
| Congenital ptosis | 4 | 10 |  |  |  | 3 | 1.39 | 2.09 |
| Congenital ptosis | 4 | 15 |  |  |  | 3 | 0.80 | 1.19 |
| Congenital ptosis | 6 | 8 |  |  |  | 2 | 2.89 | 3.97 |
| Congenital ptosis | 6 | 15 |  |  |  | 3 | 1.24 | 1.80 |
| Congenital ptosis | 7 | 15 |  |  |  | 3 | 3.71 | 6.19 |
| Congenital ptosis | 12 | 15 |  |  |  | 2 | 1.06 | 1.49 |

Abbreviations: CCDD-congenital cranial dysinnervation disorder, CFEOM-congenital fibrosis of the extraocular muscles, DRS-Duane retraction syndrome, oCCDD-ocular congenital cranial dysinnervation disorder, OEadj-adjusted observed/expected ratio, OEun-unadjusted observed/expected ratio, 1-facial paralysis, 2-hearing impairment, 3-lower cranial nerve, 4-central nervous system structural/functional malformation, 5-peripheral nervous system/muscle/connective tissue, 6-craniofacial, 7-non-craniofacial dysmorphisms, 8- skeletal (non-scoliosis), 9-skeletal (scoliosis), 10-pulmonary/ lung/ respiratory, 11-cardiovascular, 12-gastrointestinal (GI), 13- renal/urinary/genital, 14-endocrine, 15-skin/ hair/ teeth/nails.

**Supplementary Table 11.**
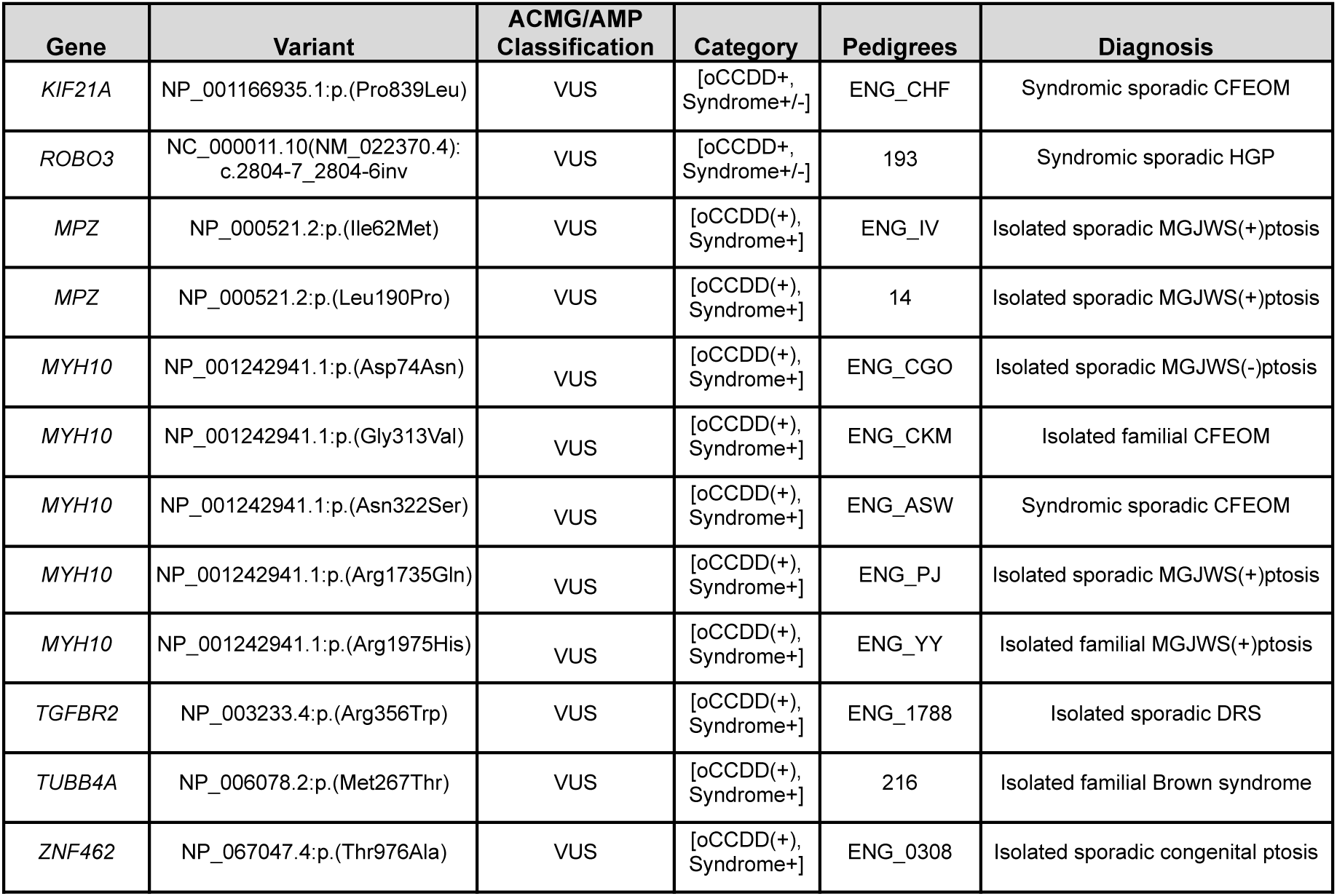
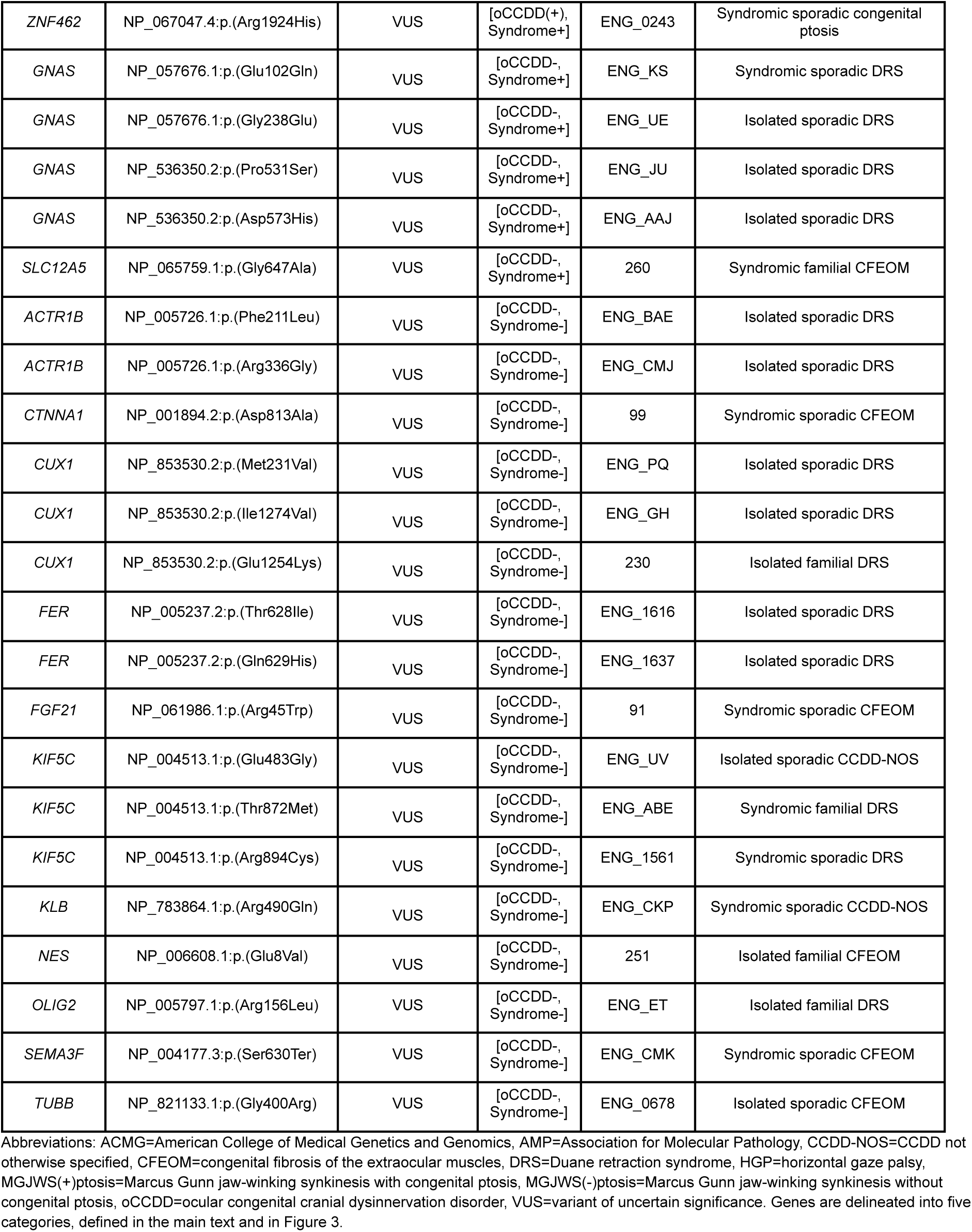
Additional novel oCCDD candidate genes/variants of uncertain significance that may merit additional study.

